# Translational bioinformatics and machine learning framework for biomarker discovery, disease prediction, and patient profiling for precision medicine

**DOI:** 10.64898/2026.05.23.26353961

**Authors:** Zeeshan Ahmed, Prithvi Govindareddy, William DeGroat, Rishabh Narayanan, Elizabeth Peker, Saman Zeeshan

**Affiliations:** Department of Medicine, Robert Wood Johnson Medical School, Rutgers Health, 125 Paterson St, New Brunswick, NJ, 08901, USA; Rutgers Institute for Health, Health Care Policy and Aging Research, Rutgers Health, 112 Paterson St, New Brunswick, 08901, NJ, USA; Department of Biomedical and Health Informatics, UMKC School of Medicine, 2411 Holmes Street, Kansas City, 64108, MO, USA

**Keywords:** Multi-omics, Genomics, Bioinformatics, Machine Learning, Biomarker, Disease Prediction, Patient Profile, Precision medicine

## Abstract

Precision medicine aims to advance our ability from a “one-size-fits-all” approach to personalized and predictive healthcare across diverse populations. It promotes integration of multi-omics and phenotypic data to understand disease mechanisms and discover novel biomarkers and risk factors, which could be used to predict and prevent critical diseases in individual patients across diverse populations. The potential implications of precision medicine approach can accelerate our ability to classify patients at higher risk of developing critical diseases, improve diagnostic capabilities, develop deeper understanding of individual risk, investigate racial differences and demographic characteristics, and find relationships between genetic variants, expressions, and diseases. This study focuses on implementing an innovative and data driven framework of translational bioinformatics and Machine Learning (ML) techniques to analyze multi-omics, including RNA-seq and Whole-Genome Sequencing (WGS) data, generated using blood samples of randomly consented patients. First, we utilized bioinformatics pipelines to identify differentially expressed genes and their pathogenic and likely pathogenic variants for the downstream data analysis, annotation, and visualization. Then, applied a nexus of ML models for multi-omics biomarker discovery, disease prediction, density-based clustering, single-patient profiling, and pathogenicity classification. WGS data analysis supported the exploration of genetic variation and diversity among patients to identify known and novel biomarkers, whereas RNA-seq data analysis improved our understanding of functional and biological pathways that underlying disease states. We classified and clustered pathogenic variants and expressions across various genes and discovered numerous diseases leading risk factors. Our results include gene-disease associations and captured common pathways across the broader population, demonstrating a level of sensitivity and accuracy that has broad clinical implications. We validated our results through clinical records, and state of the science literature. This study delves into the strengths of multi-omics data integration and capabilities of ML application in genetically diverse and complex patient cohorts. Our approach has the potential to elucidate complex gene-disease interactions for genetically diverse populations, which can support earlier diagnoses for patients in many disease realms.

## Introduction

The prevalence of diagnostic errors in medicine result in substantial but preventable harm across various medical fields [1, 2, 3, 4]. This is largely because most modern medicine depends on the treatment of generic symptoms and empirical evidence from clinical trials rather than focusing on the personalized care tailored to unique biological composition of individual patients [5, 6]. Critically, reducing the rate of misdiagnosis will ultimately improve patient outcomes, increase available treatment options, and address growing costs. The field of precision medicine has gained increasing attention for its potential to transform patient care through individualized diagnosis and treatment strategies. To determine key biological indicators that distinguish healthy and diseased patients, it is imperative to comprehend the particularities and unique aspects of each patient [7]. In aspiring to improve diagnostic accuracy by understanding the intricacies of human biology, scientists frequently explore various types of omics, which include e.g., genomics, transcriptomics, epigenomics, proteomics, microbiomics, metabolomics, and multimodal integrations of those [8, 9, 10, 11]. In clinical settings, the timely identification of significant patterns among millions of these multi-omics and clinical features is paramount for discovering underlying biological pathways and modifiable risk factors to support early detection and prevention of complex and rare disorders e.g., neural, respiratory, cardiovascular, immunological, infectious, cancer and skeletal [12]. However, individual omics modalities alone are insufficient to capture the full complexity of biological systems [13]. The effective integration of multi-omics with phenotypic data has the potential to generate holistic multimodal patient profiles, necessary to underscore key characteristics and improve prognosis, support early diagnosis, and provide more economical personalized treatment [7].

Traditional bioinformatics and statistical modelling have long been the default choice for analyzing and interpreting big data [14]. Most importantly, Genome-wide Association Studies (GWAS) and Gene Expression Analysis (GEA) have remarkably assisted in understanding the genetic basis of human disease by uncovering millions of loci associated with various complex phenotypes [15, 16, 17]. Whole-Genome Sequencing (WGS) data analysis supported the exploration of genetic variation and diversity among patients to identify known and novel biomarkers [18], whereas RNA-seq data analysis improved our understanding of functional and biological pathways that underlie disease states [19]. Although these approaches provide important insights into multifactorial diseases, they have limitations when used individually. RNA-seq based gene expression analysis on its own fails to capture the effects of non-coding genes, whereas linking WGS variant data to specific diseases remains challenging [19]. Most genetic studies focus on a targeted set of disease-specific genes, rather than an unbiased investigation of the entire human genome and transcriptome, limiting the applicability and generalizability of the results in clinical settings. Furthermore, these are unable to predict disease and detect all the heritability explained by Single Nucleotide Polymorphisms (SNPs) and can only target specific variants of complex diseases [18]. The unbiased integration of comprehensive multi-omics data can provide a deeper view into causal relationships between genetic variants, expressions, and complex diseases. However, it remains persistently difficult due to the inherent heterogeneity and large volumes of multi-omics data [20]. Progress in the development of translational bioinformatic approaches has significantly enabled robust data processing and investigation of high-throughput omics data [20]. However, due to their inability to integrate heterogeneous data types, these tools have historically underperformed in identifying multi-faceted interactions in high-dimensional multi-omics data [21].

The cutting-edge Artificial Intelligence (AI) and Machine Learning (ML) approaches have proven effective at uncovering elucidative knowledge on disease-causing biomarkers and the biological underpinnings of a plethora of human diseases [7]. The analysis of integrated multi-omics and phenotypic data with a nexus of translational bioinformatics and ML techniques has the potential to accurately predict disease risk in patients by identifying patterns that may otherwise be eluded by the traditional bioinformatics approaches [22]. Furthermore, the current state-of-science ML techniques typically involve employing single-model supervised ML on pre-labelled case/control cohorts, have proven less effective at disease prediction, especially when using multi-omics data [21, 23]. In response, we developed multiple ML pipelines and applications that support flexible and ensemble-based analytics frameworks, capable of predicting diseases with higher accuracy in a case/control cohort [23, 24, 25, 26, 27]. However, to achieve unbiased and effective personalized treatment, it is imperative to equally explore and develop unsupervised learning techniques on patients whose disease states are unknown a priori. The current unsupervised ML methods remain substantially less developed than supervised approaches, often requiring extensive amounts of prelabelled data [28, 29]. Since unsupervised techniques involve finding associations among data points to describe higher-level patterns, the importance of unbiased representation within a population is paramount [30].

In this study, we implemented a FAIR (Findable, Accessible, Interoperable, Reusable) and data driven nexus of translational bioinformatics and ML techniques to analyze unlabeled multi-omics data from a cohort of randomly consented patients. Random participant selection rather than disease-specific targeted recruitment was planned and helped preserve genetic diversity that may better guide unsupervised clustering and discriminatory ML models. Genomic and transcriptomic data generated from the collected blood sample of each patient were independently analyzed and reintegrated to identify predictive biomarkers and associated disease states. Nevertheless, this integration remains challenging due to the heterogeneity and scale of multi-omics data.

## Results

The reported results are achieved by analyzing multi-omics data (RNA-seq and WGS, generated from blood samples) of randomly consented patients (n=96) with the implementation of designed translational bioinformatics and data driven ML framework, explained in the methods section (Figure 1). These results include classified, clustered, and integrated gene expressions and variants, annotated gene-disease associations, and common pathways to understand relationships between disease causing multi-omics biomarkers and clinical outcomes. Furthermore, we produced single-patient profiles based on the discovered biomarkers and associated diseases, and presented overall disease distribution across patient cohort, including the most commonly identified biomarkers and associated diseases.

**Figure 1.**
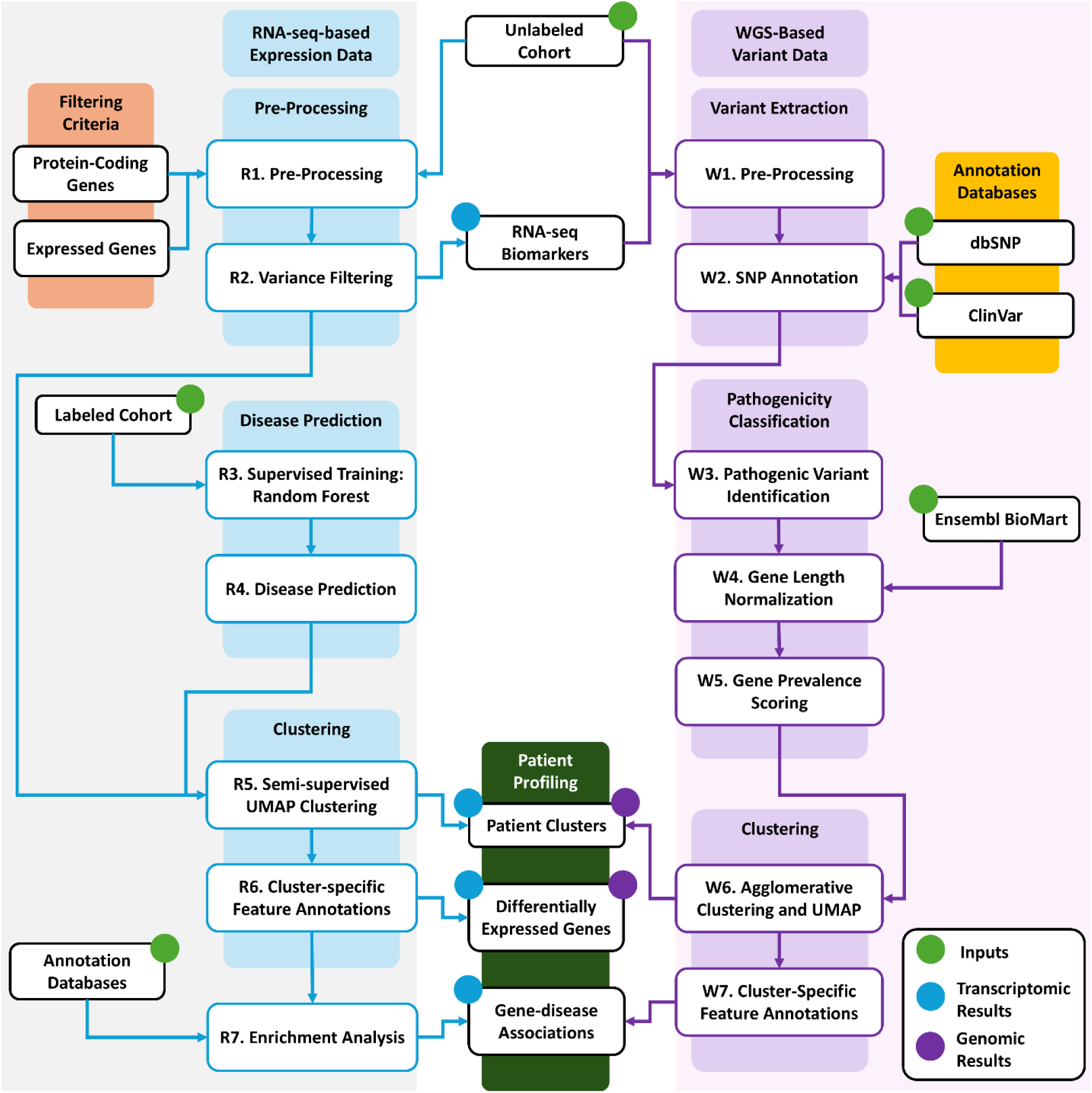
Data analysis methodology, divided into two sections. RNA-seq driven expression data analysis and WGS based variant data analysis.

### Gene expression analysis-based results

Gene expression analysis revealed 11,235 protein-coding genes from 56,861 transcripts with a mean TPM greater than 10 thresholds. Expression was visualized by generating heatmap across the cohort (Figure 2A, 2B, and 2C). Using a nested cross-validation scheme, our calibrated disease prediction pipeline achieved a mean F1 score (*pos label* = 0) of 0.672 (95% CI = 0.580, 0.752) and an MCC of 0.649 (95% CI = 0.550, 0.738). The model attained a balanced accuracy of 0.810 (95% CI = 0.755, 0.862) and an overall accuracy of 0.912 (95% CI = 0.892, 0.933). We explored how classification thresholds affect the model’s performance and varied the decision threshold from 0.1 to 0.9. F1 scores ranged from 0.190 to 0.700 and balanced accuracy from 0.551 to 0.844, illustrating the trade-off between precision and recall at different probabilities. A threshold of 0.70 produced the highest F1 (0.700) without sacrificing balanced accuracy (0.844). This was used to make “confident” positive predictions. We utilized the mRMR algorithm with an MIQ criterion to select genes (n=15) representative of the Cardiovascular Disease (CVD) classification: *GPSM3*, *RPS15*, *GSTO1*, *GSTK1*, *GPX4*, *RPS11*, *RPLP2*, *RPS14*, *RPL37A*, *GMFG*, *RPS18*, *RPS19BP1*, *GABARAP*, *RPS10*, and *RPS21*. Before applying the model, outlier samples (SID = 666, 675, 693, 694, 698, 711, 731, 732, and 743) were removed based on PCA criterion of two standard deviations from the center of the distribution. With that, we predicted occurrence of CVD in 30 patients.

**Figure 2.**
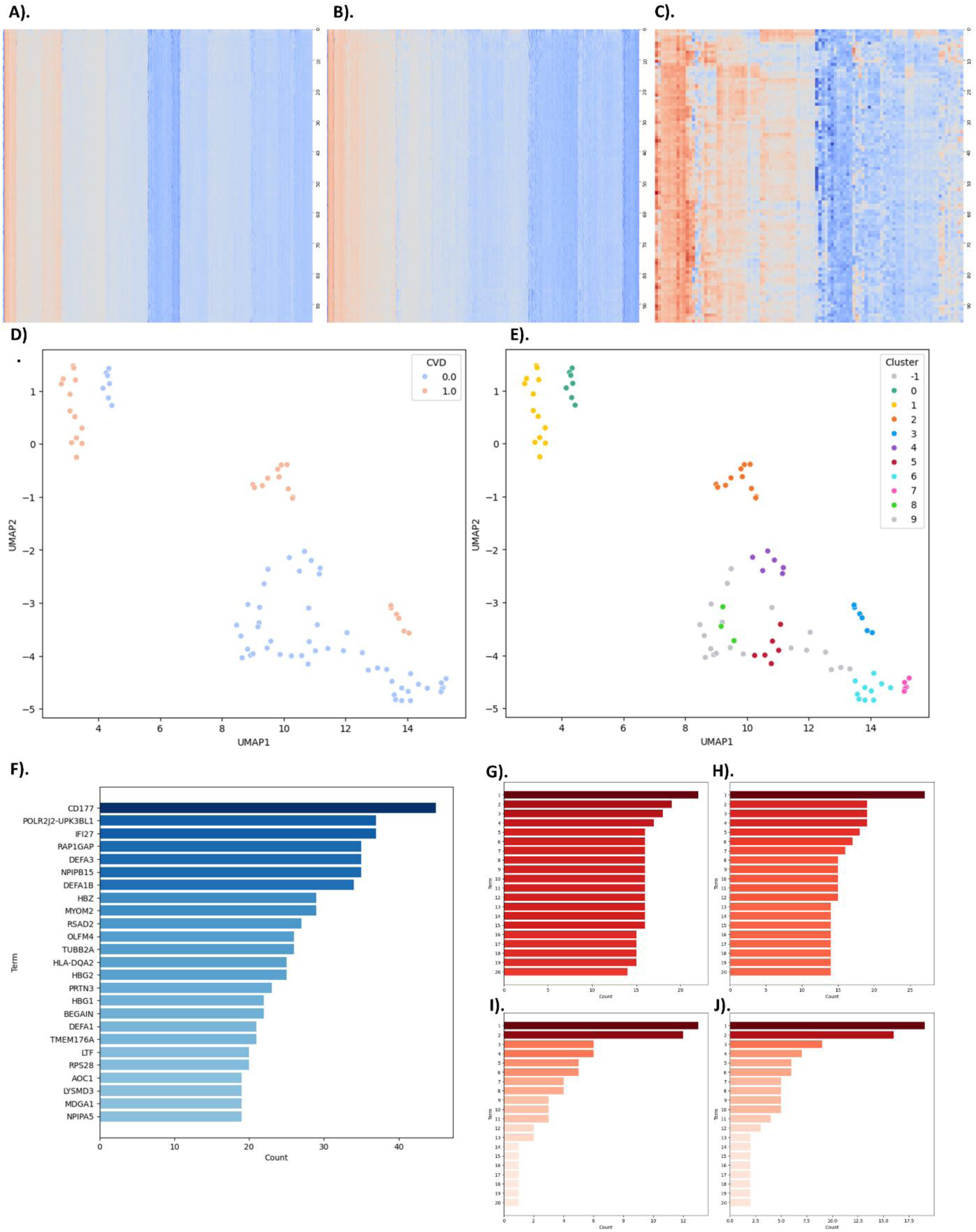
RNA-seq driven gene-expression data analysis results. **(A)** Heatmap containing all genes that passed RNA-seq preprocessing. **(B)** Heatmap of 5,000 variance-filtered genes utilized in clustering analysis. **(C)** Heatmap of 100 variance-filtered genes. **(D)** UMAP embedding shows patients predicted to have CVD. **(E)** UMAP embedding showing HDBSCAN-generated clusters of patients. **(F)** Most occurrent genes in RNA-seq-based subject-specific profiles. **(G)** Most frequent annotations in GWAS Catalog. **(H)** Most frequent annotations in DisGeNET. **(I)** Most frequent annotations in OMIM Disease. **(J)** Most frequent annotations in ClinVar.

UMAP, followed by HDBSCAN clustering, identified ten clusters in the pseudo-labeled dataset, along with a set of outlier samples that did not fit any cluster’s criteria. Clusters 1, 2, and 3 overlapped with the patients predicted to have CVD (Figure 2D, 2E), suggesting a clear separation of disease-associated transcriptional profiles. DE and subsequent enrichment analysis indicated that three of the clusters (1, 6, and 7) were particularly enriched for disease-associated biological pathways. Using our individualized KNN-weight reference approach, each patient was compared against a smooth, population-level baseline and ranked according to the twenty genes with the largest absolute LFC values. Patient-specific enrichment analysis of these twenty genes against GWAS catalog, ClinVar, DisGeNET, and Online Mendelian Inheritance in Man (OMIM) Disease highlighted recurrent disease-relevant terms and pathways across the cohort. *CD177* appeared frequently among the top genes (Figure 2F). In GWAS catalog, the most identified terms were labelled as “Abdominal Aortic Calcification Levels”, “*CD177* Antigen Levels”, and “Response to SSRI (Symptom Remission)” (Figure 2G). OMIM disease annotations repeatedly included “blood”, “ichthyosis”, and “lymphoma” (Figure 2H). DisGeNET terms focused on “Myeloid Leukemia, Chronic”, “Thalassemia”, and “Inflammation” (Figure 2I). Finally, ClinVar analysis frequently identified “Diamond-Blackfan anemia”, “severe congenital neutropenia”, and “susceptibility to malaria” (Figure 2J). Collectively, these results underscore the capacity of this pipeline to reveal individualized disease associations while also capturing common pathways across the broader population.

### Gene variant analysis-based results

Mutation profiling revealed 43,451,462 from a total of 396,788,923 variants across all 96 patients of our cohort for the top 5000 DEG derived from RNA-seq expression analysis. Annotations were sourced from dbSNP and ClinVar to provide crucial metadata for each variant, including known RS numbers, clinical significance, and associated clinical disease names. Further preprocessing was performed to identify 972,823 variants with known RS numbers, constituting approximately 0.25% of our original dataset and serving as the basis for subsequent pathogenicity classification, clustering, and visualization. The identification of pathogenic, likely pathogenic, and risk-factor variants was supported through ClinVar disease annotations. A large majority of the filtered variants were marked benign or of unknown pathogenicity and consequently pruned. Only 1,747 total variants (0.18% of the filtered set) across 84 genes were identified as pathogenic or known risk factors for diseases, with between 10 and 26 variants recalled for each patient in our cohort. Normalized counts for these 84 genes are displayed on a logarithmic scale in Figure 3A. Gene disease associations were identified by cross-referencing gene information from dbSNP with disease metadata from ClinVar. We discovered 241 different diseases associated with these 1,747 pathogenic variants. Some of the most common diseases associated with patients in our cohort included myocardial infarction, tuberculosis, malaria, bacteremia, asthma, and diabetes. Figure 3B visualizes the distribution of all 241 identified diseases, while Figure 3C depicts the 50 most frequent diseases among all participants. Approximately 37% (88 diseases) of all 241 identified diseases were predicted for only one patient, while only 20% (49 diseases) were predicted in at least 10 patients. This relative sparsity of shared variants and diseases is attributable to the high genetic diversity of our random cohort. Agglomerative hierarchical clustering was performed using a matrix of variant prevalence to identify 10 distinct clusters of individuals. Visualization of the high-dimensional data necessitates lower-dimensional embedding, which was enabled through a 2D UMAP projection, as presented in Figure 3D. Separate comparative gene-disease networks for the top 50 diseases identified through WGS analysis as well as ClinVar-based diseases identified from RNA-seq analysis are presented in Figure 3E and 3F, respectively.

**Figure 3.**
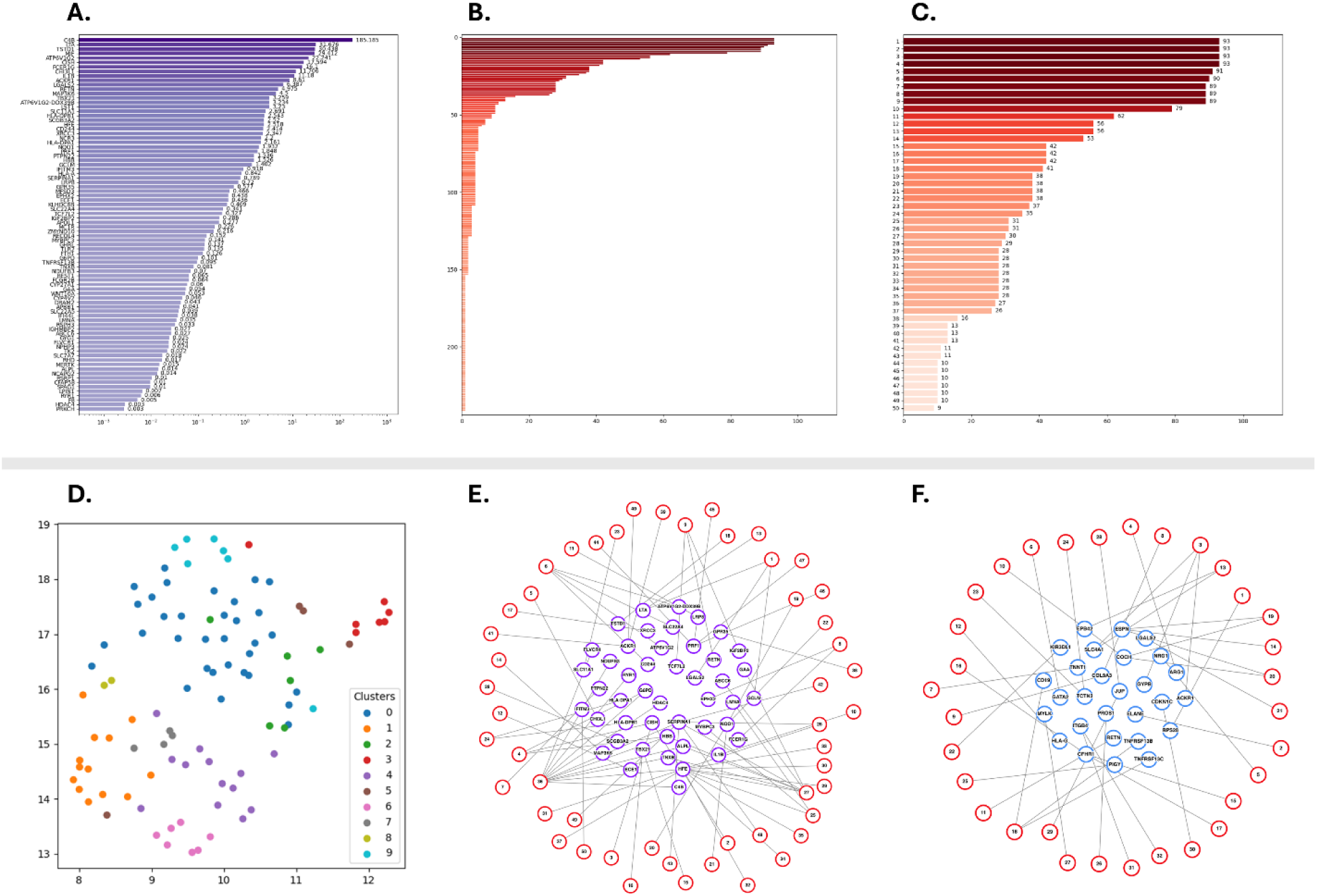
WGS based gene-variant data analysis results. **(A)** Normalized gene counts. **(B)** Disease prevalence counts. **(C)** Top 50 Disease counts. **(D)** UMAP clustering. **(E)** WGS-based gene-disease network. **(F)** RNA-seq based gene-disease network.

### Patient profiling and disease distribution across cohort

Notably, the clusters remained isolated with minimal overlap between groups. Adjacent data points are generally indicative of greater similarity between the corresponding individuals, offering greater comparability among otherwise unrelated individuals. Upon identifying gene expressions, pathogenic variants, and predicted clusters, we constructed individual profiles for each patient, including relevant biomarkers and associated diseases (Table 1 and Figure 4). Key findings include most common, rare, and complex diseases across the patient cohort (n=96, patients), divided into seventeen out of eighteen targeted disease categories described in the methods section (Figure 4A and 4B). Our analysis revealed hematologic and immune-related disorders were the most common findings, including blood and blood-forming organ diseases (81/96, patients) and immune system diseases (52/96, patients). Additionally, most influencing diseases are related to nervous system (63/96, patients); genitourinary system (51/96, patients); endocrine, nutritional and metabolic (47/96); and congenital and genetic (44/96, patients) disorders. Diseases observed with moderate occurrence in the overall cohort were related to the skin and subcutaneous tissue (36/96, patients); musculoskeletal (32/96, patients); respiratory system (32/96, patients); viral/bacterial/infectious (27/96, patients); circulatory system and CVD (20/96, patients); mental/behavioral/neurodevelopmental (18/96, patients); eye and adnexa (16/96, patients); digestive system (14/96, patients); and ear, nose and throat (13/96, patients) diseases. We discovered cancer (10/96, patients) among the complex and rare disorders, and observed some abnormal clinical findings (34/96, patients) as well (Figure 4C). The overall cohort demonstrated presence of multisystem comorbidities, excluding injuries and wounds.

**Figure 4.**
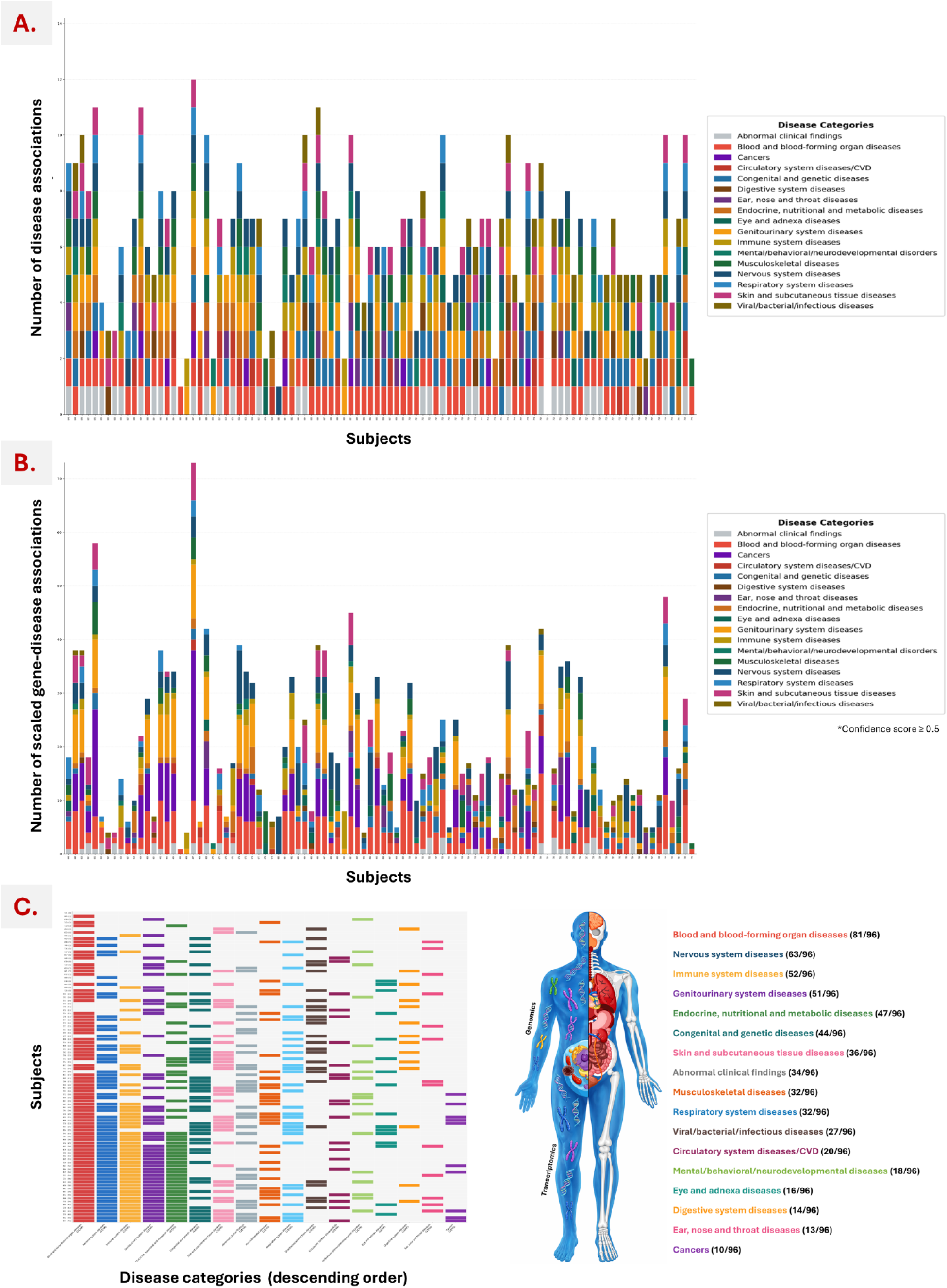
Patient profiles, gene-disease associations. A) number of diseases in patients, B) number of scaled gene-diseases associations in descending order, and C) disease distribution across patient cohort. Each bar 4 represents individual patient (A and B), whereas each color box represents its association with a disease (A, B, and C).

**Table 1.**
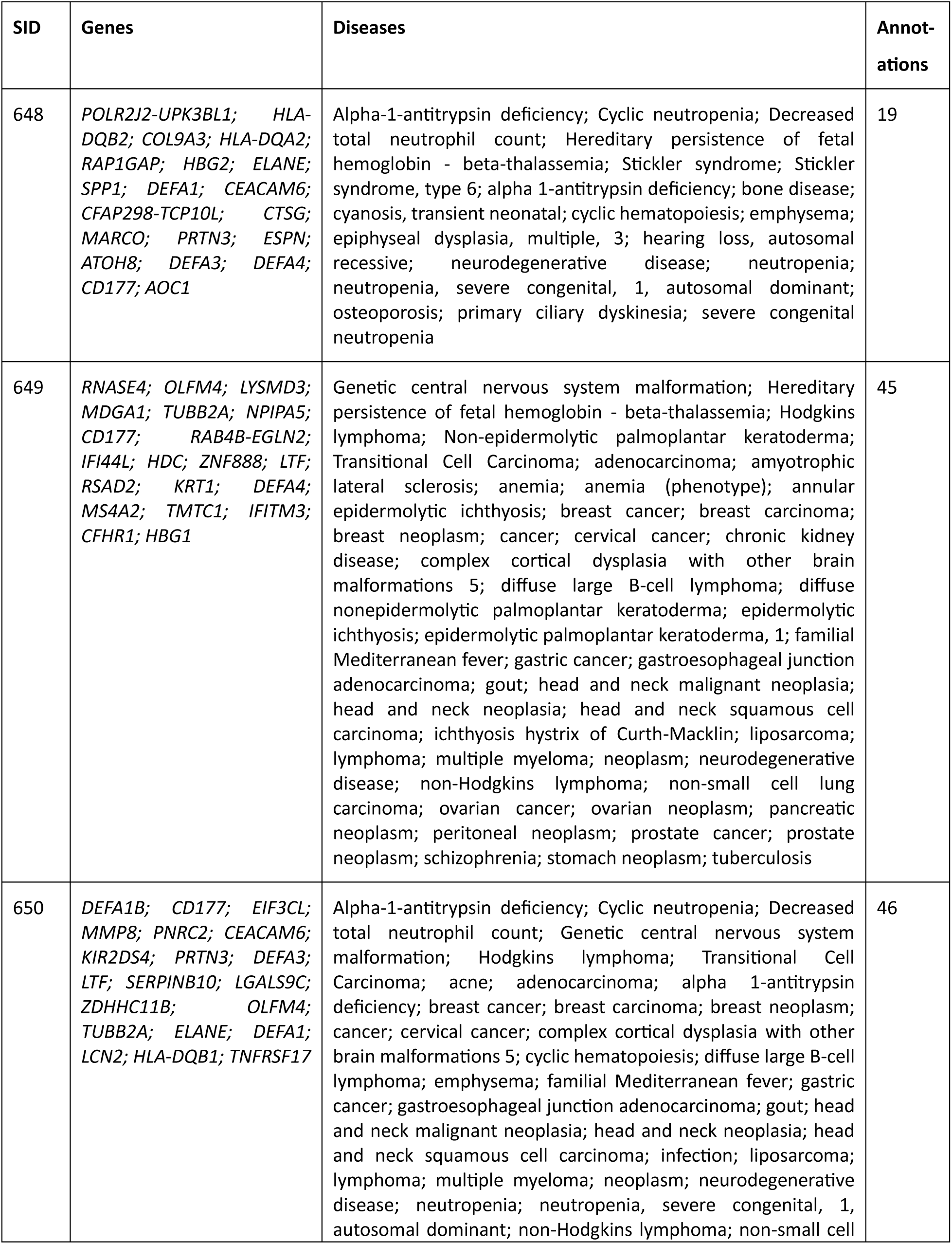
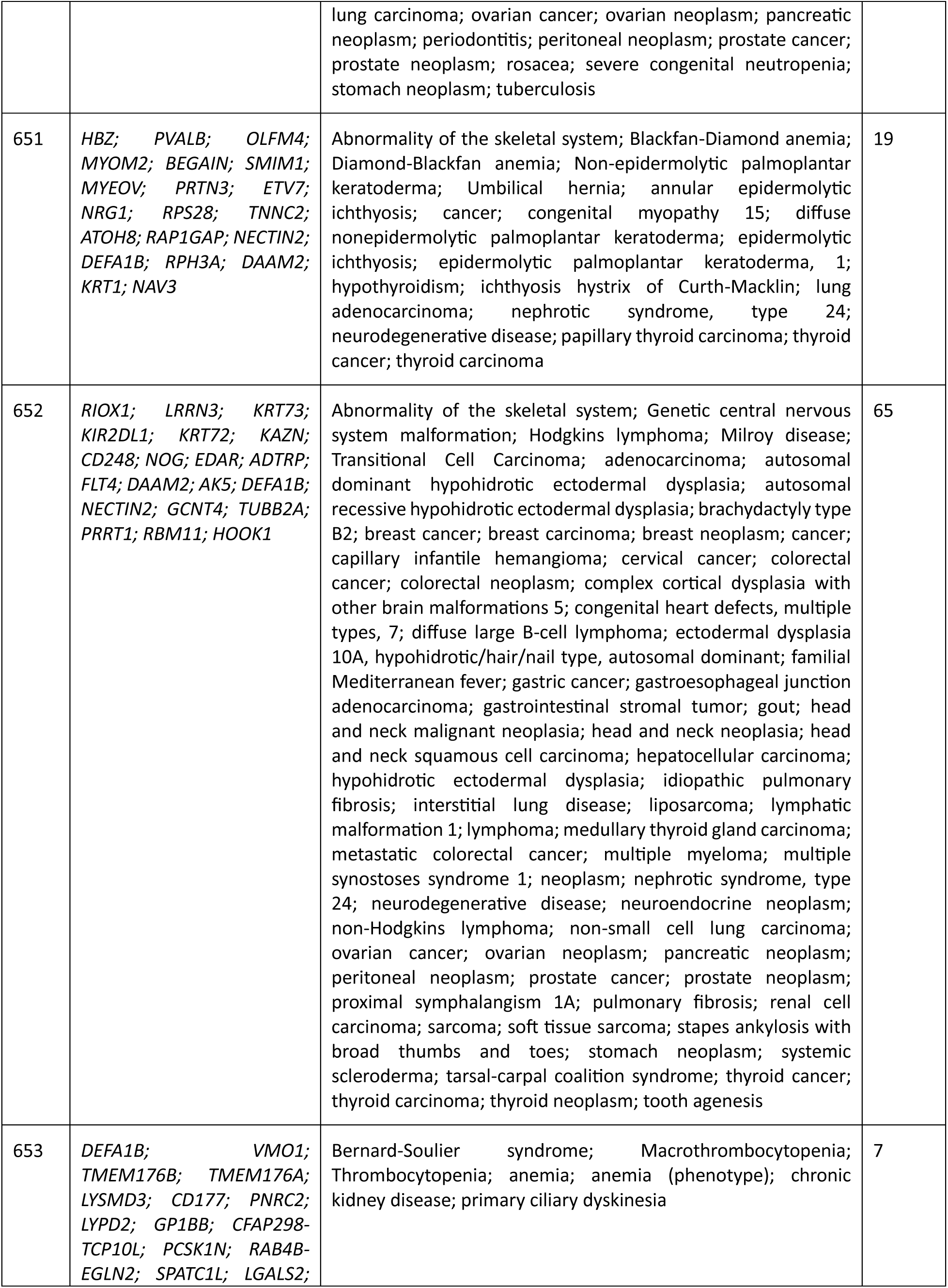
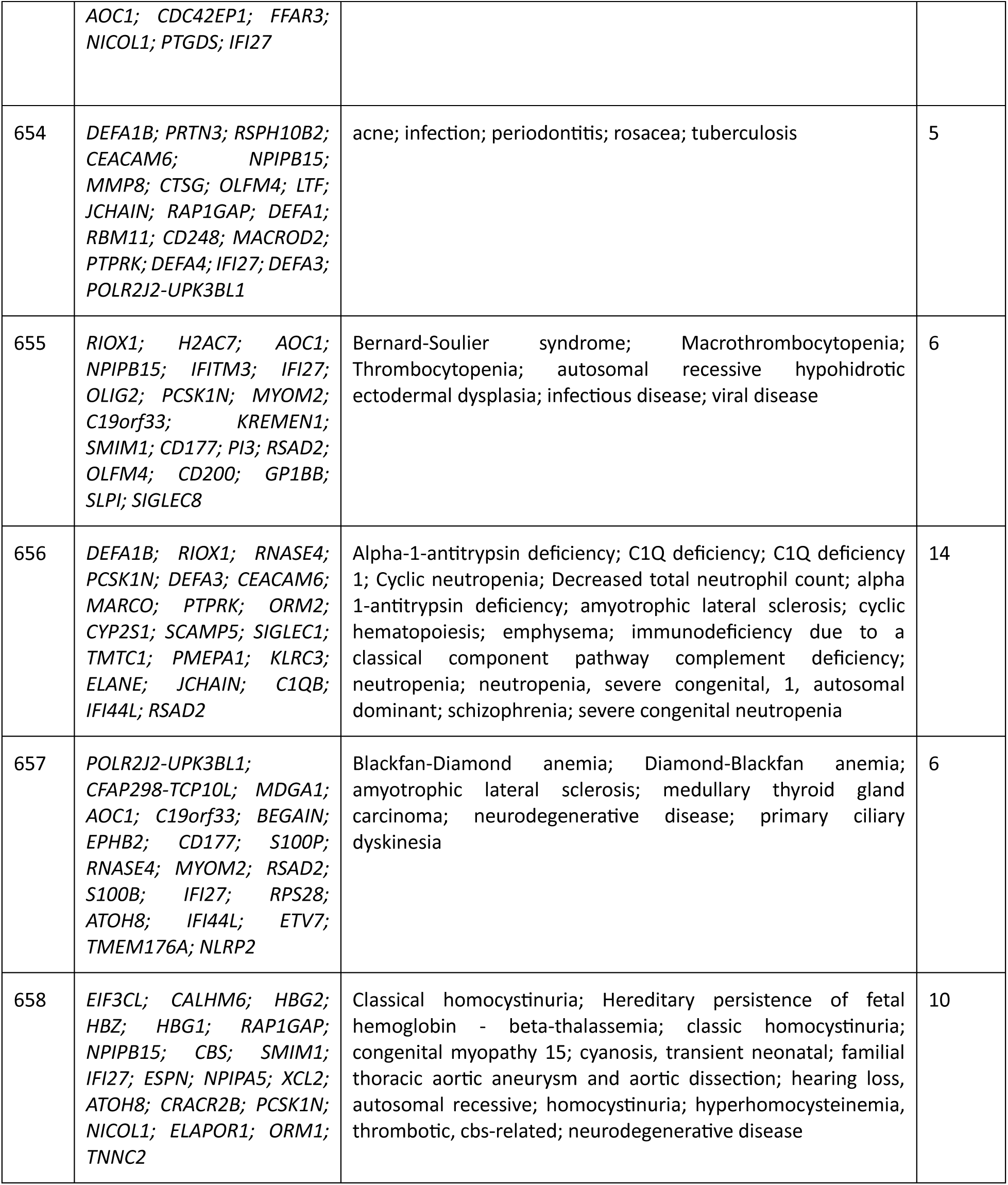
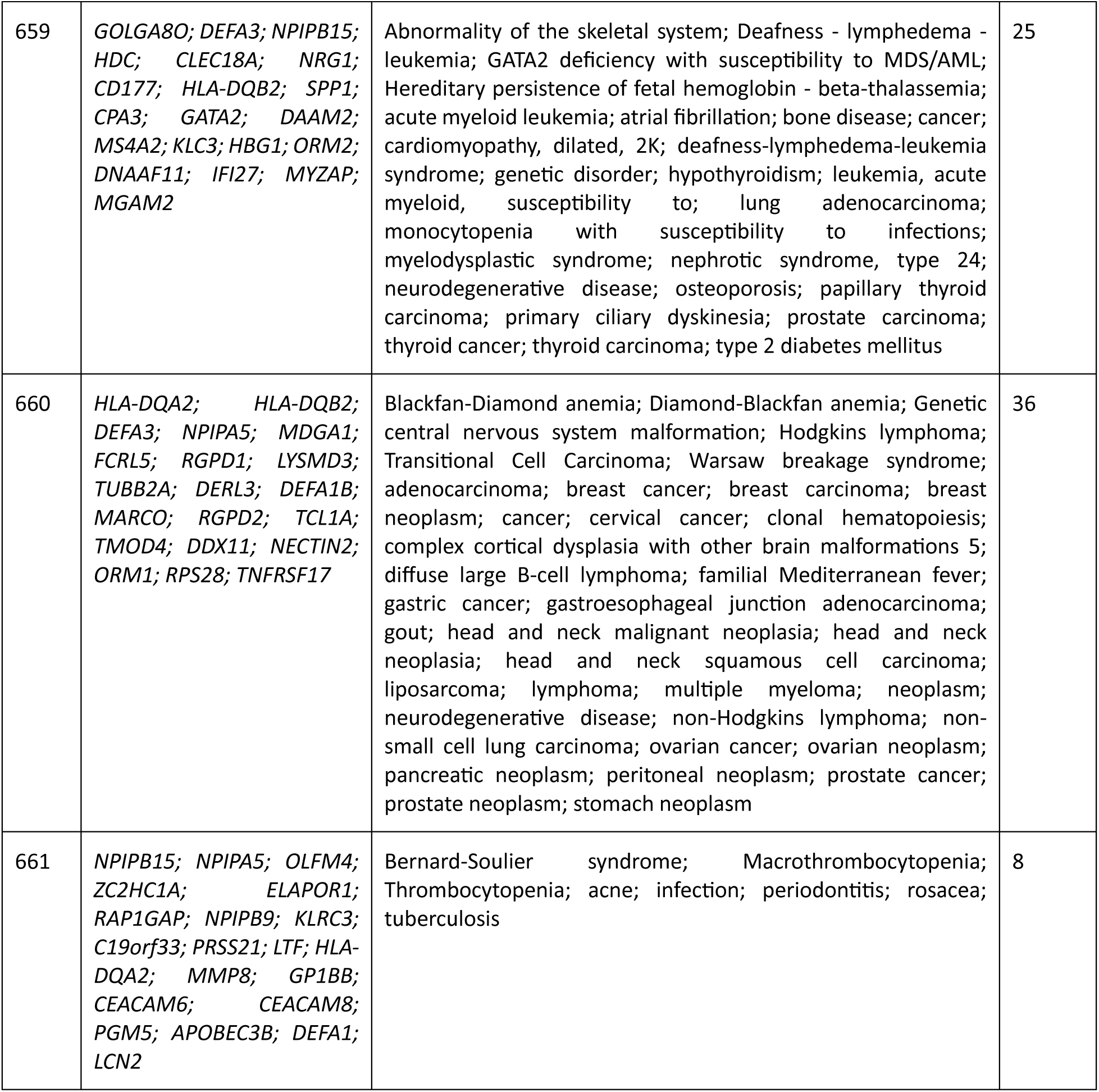
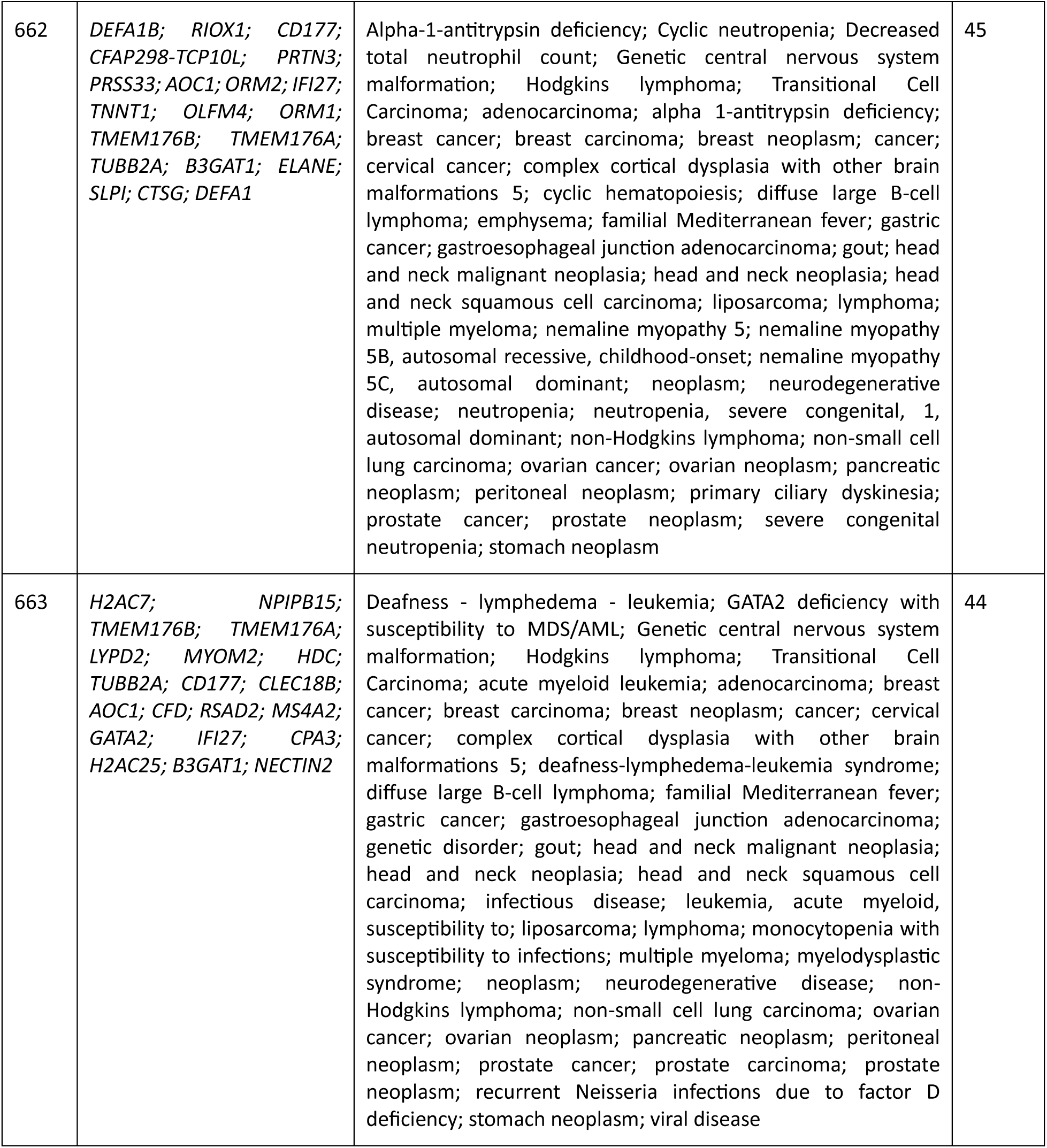
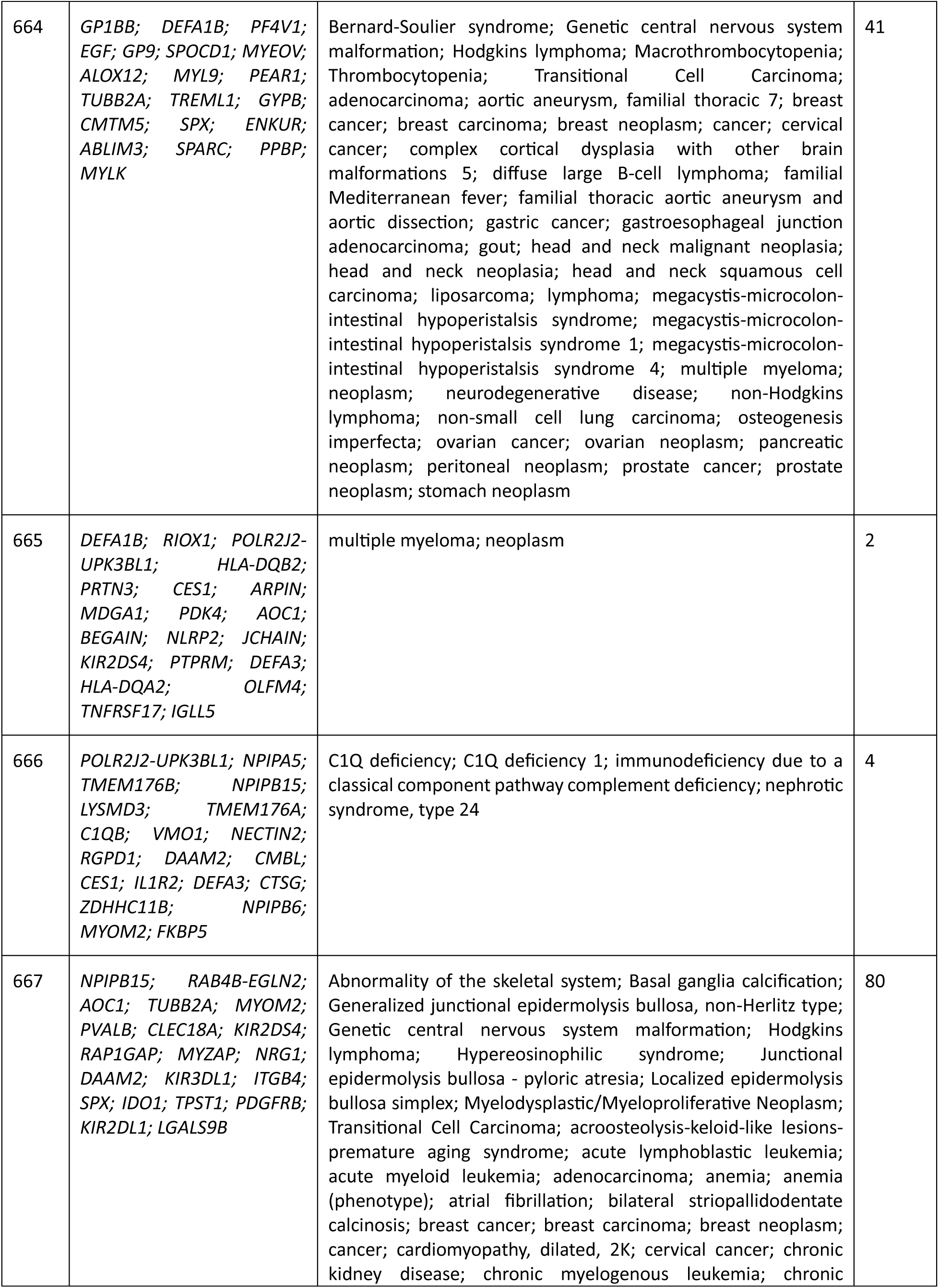
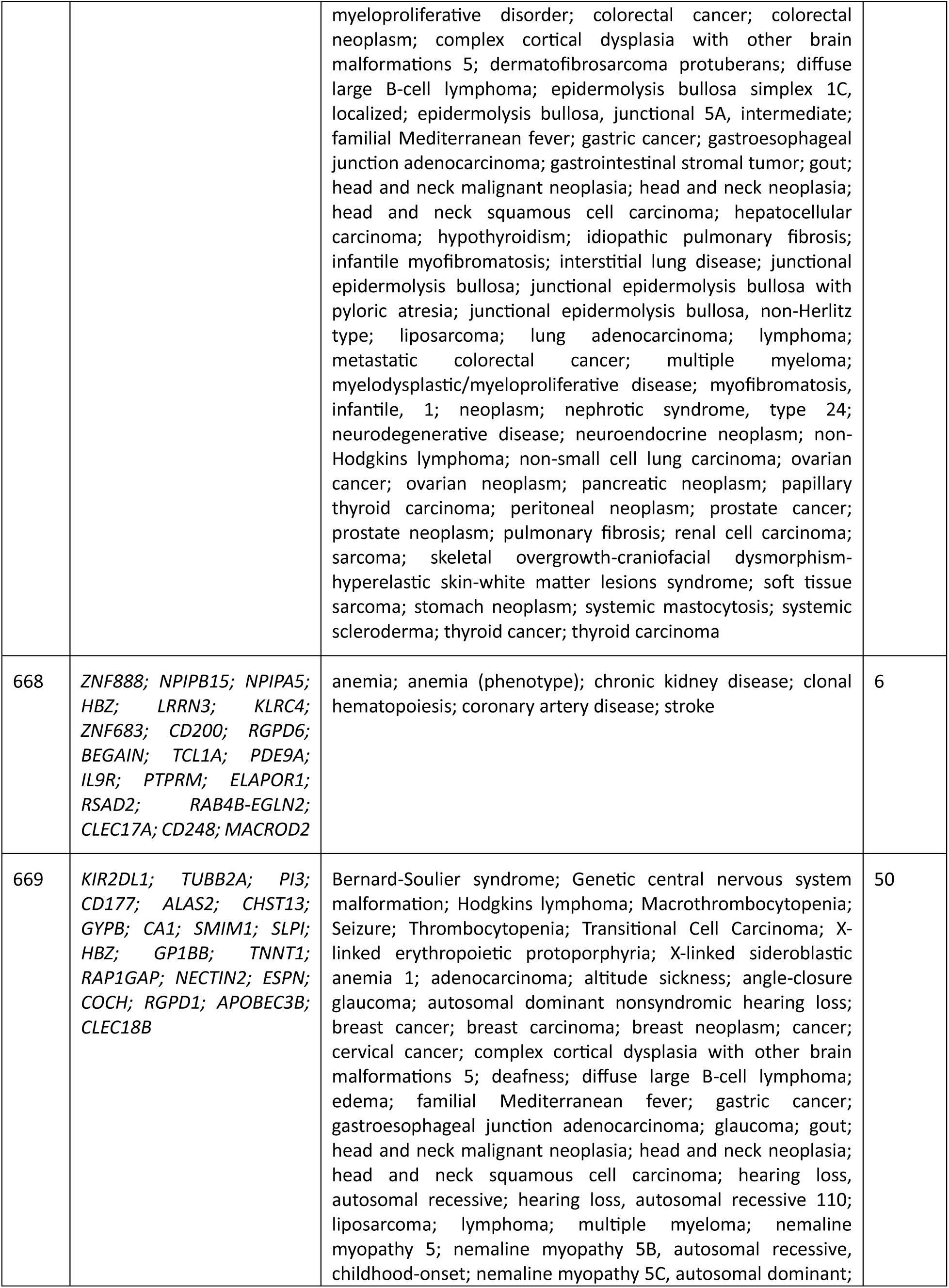
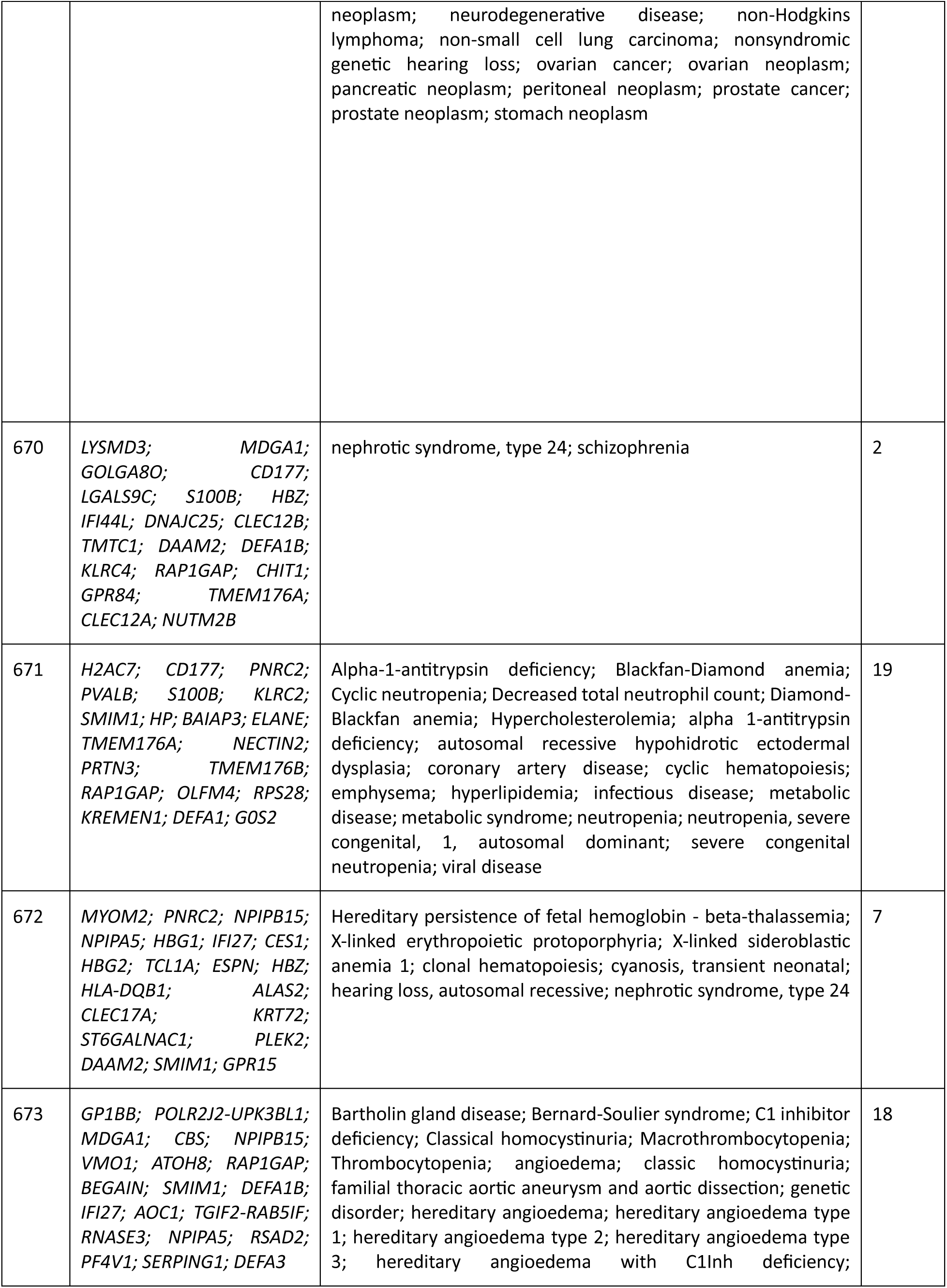
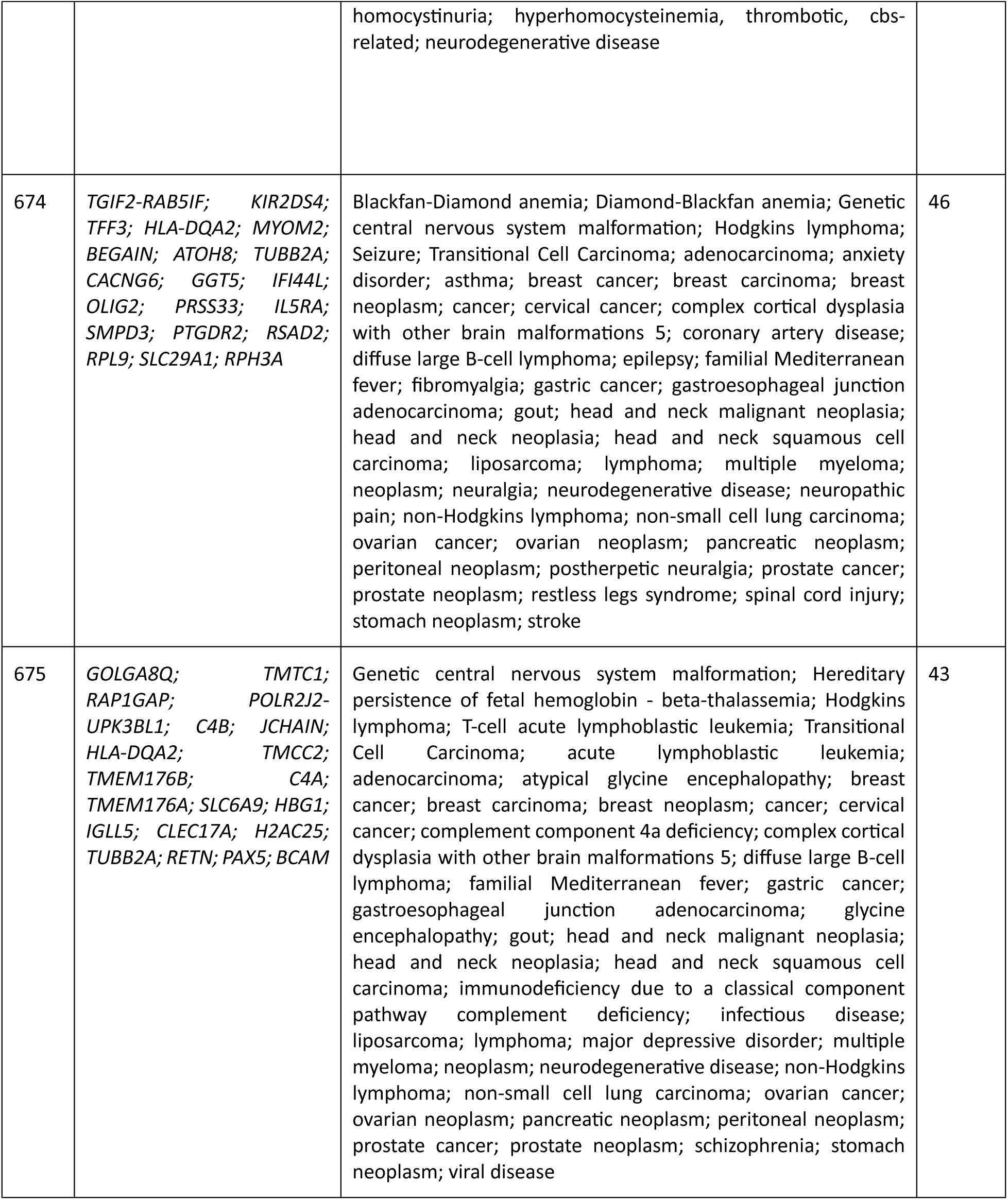
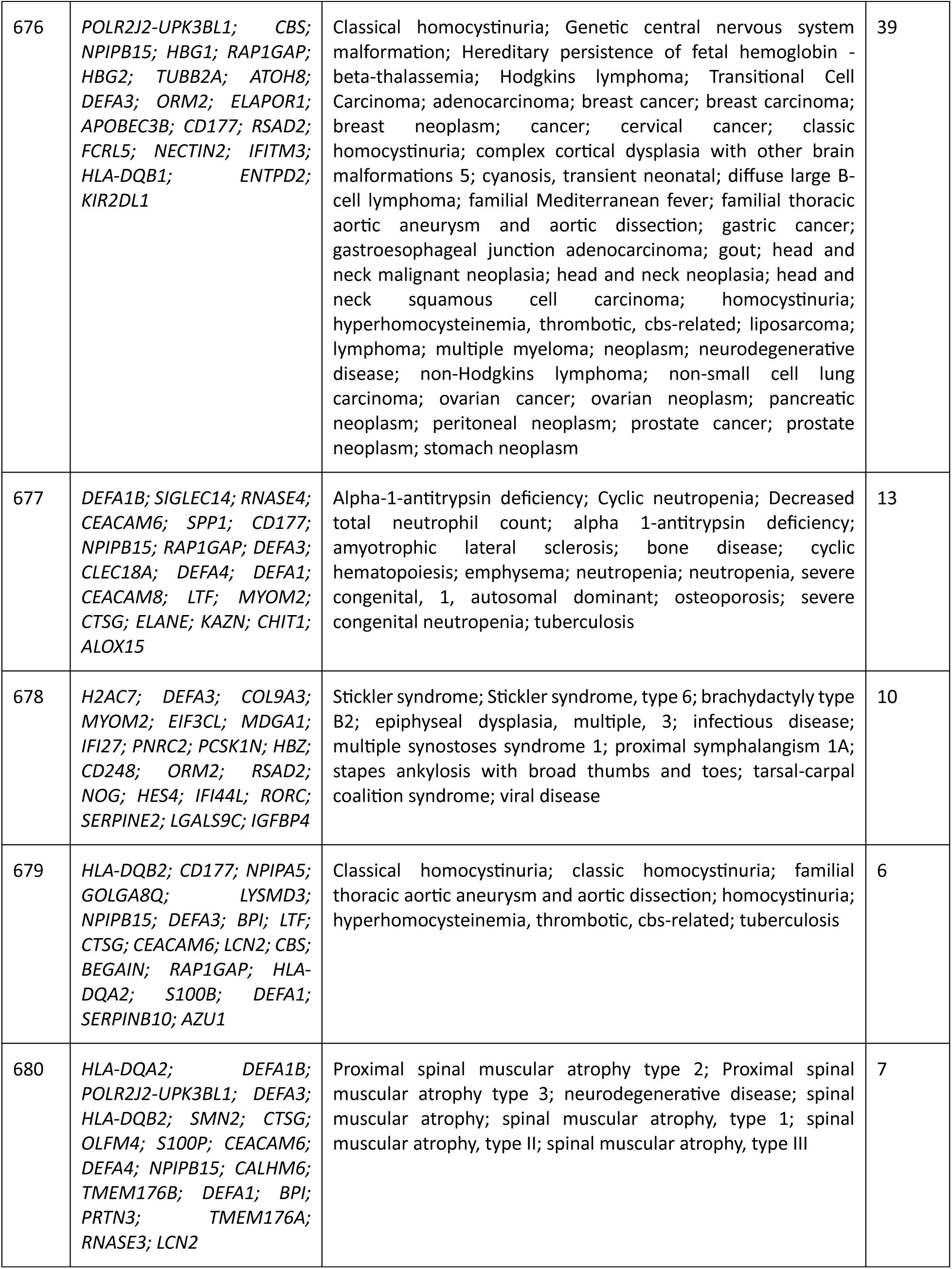
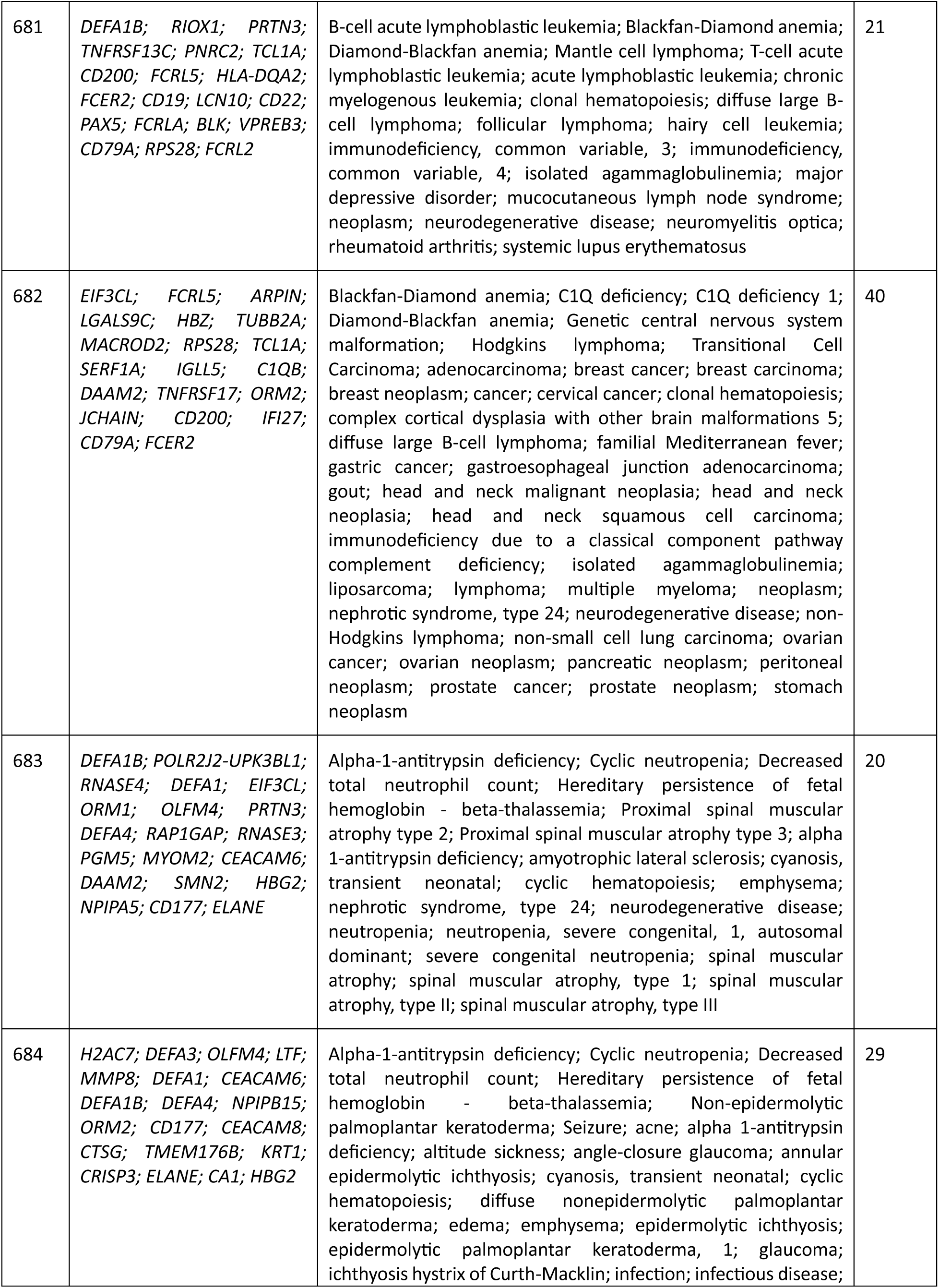
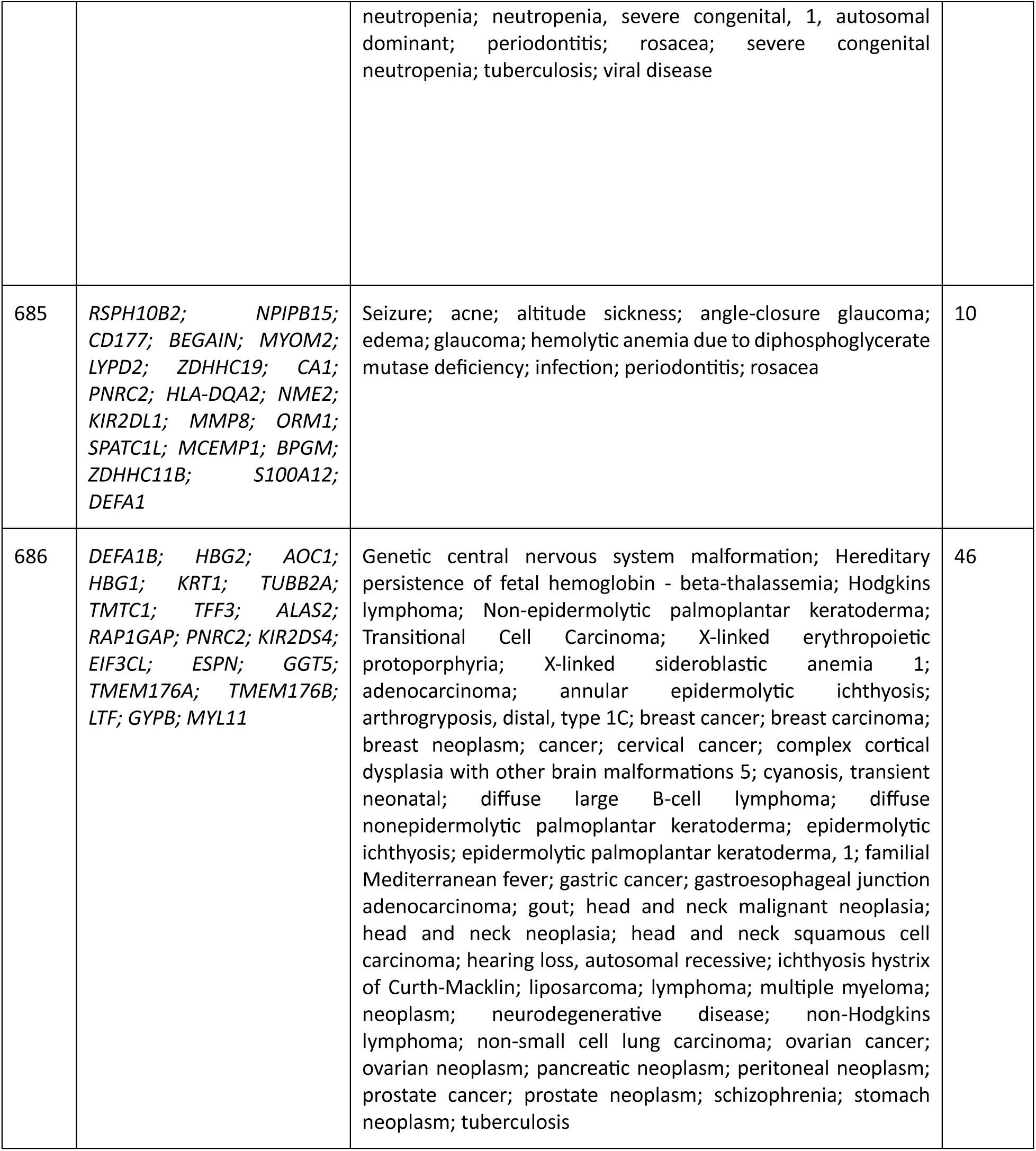
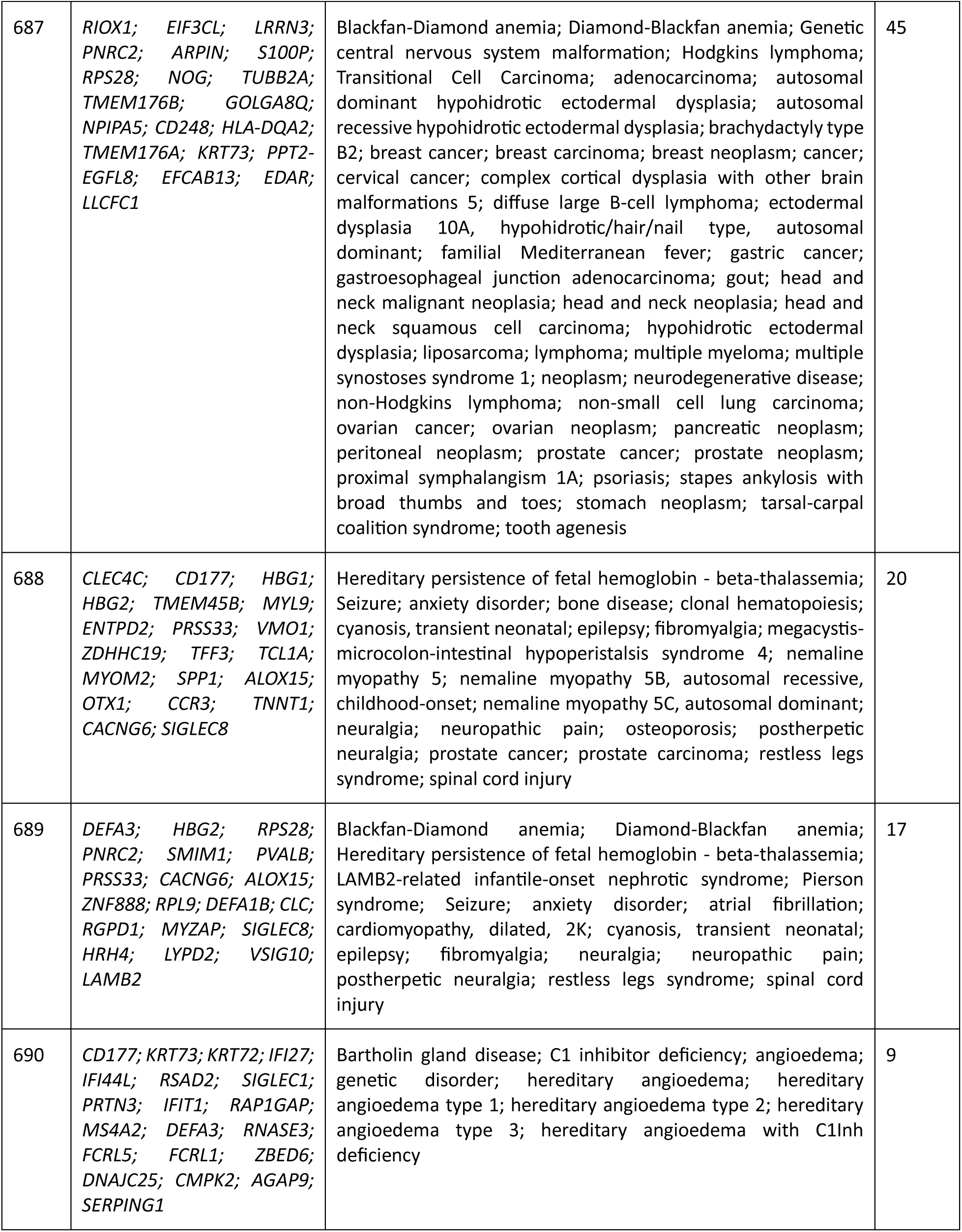
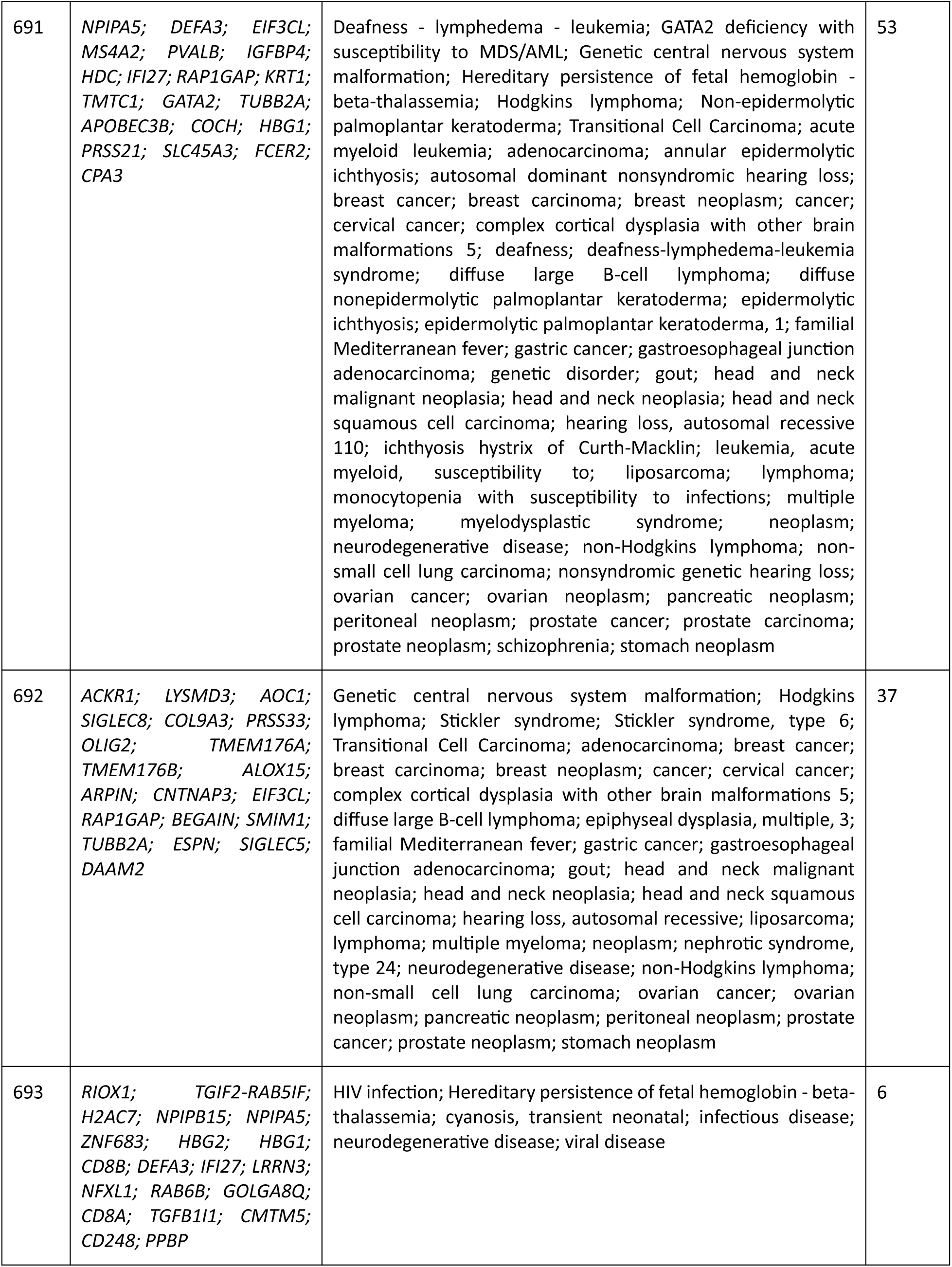
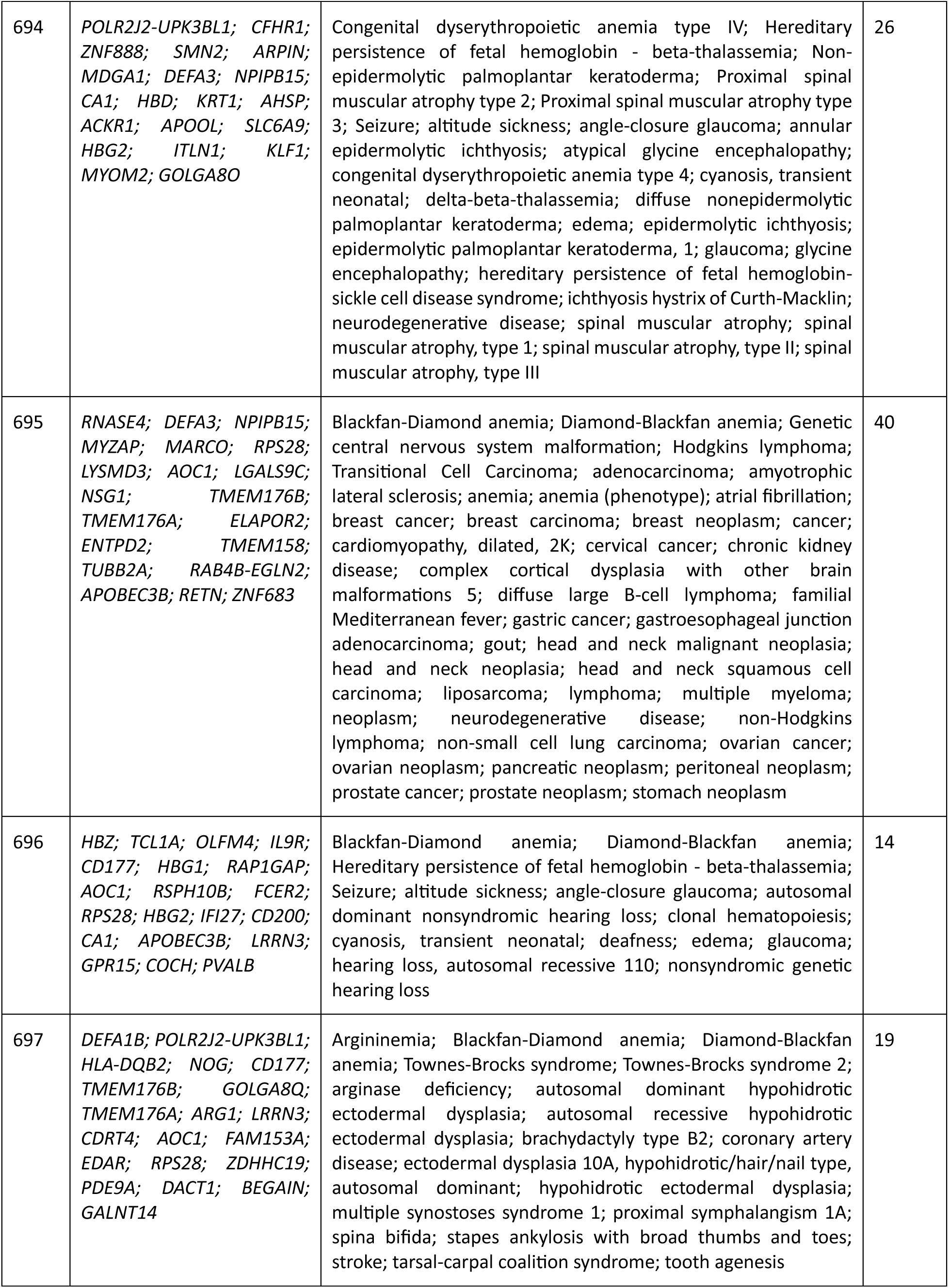
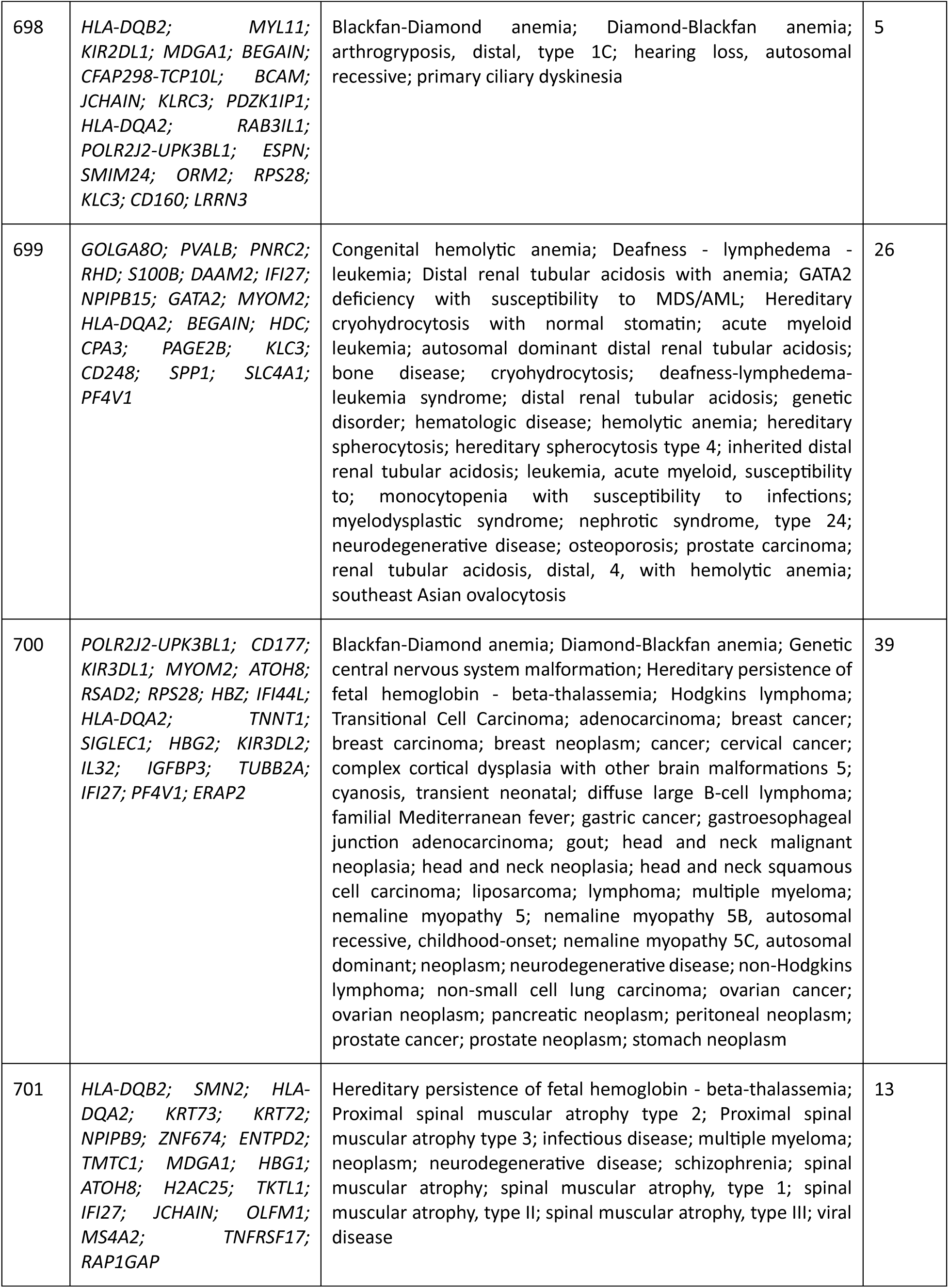
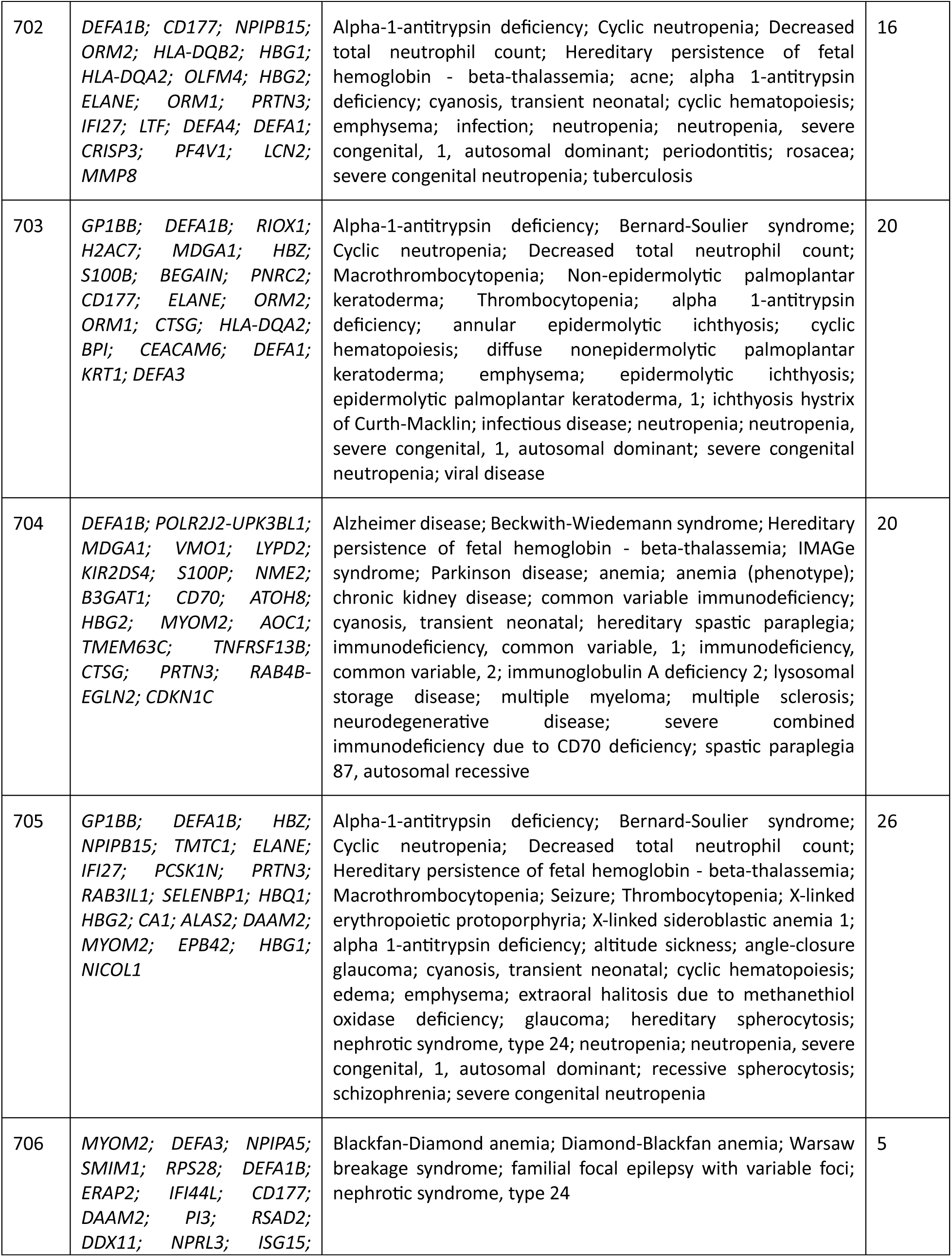
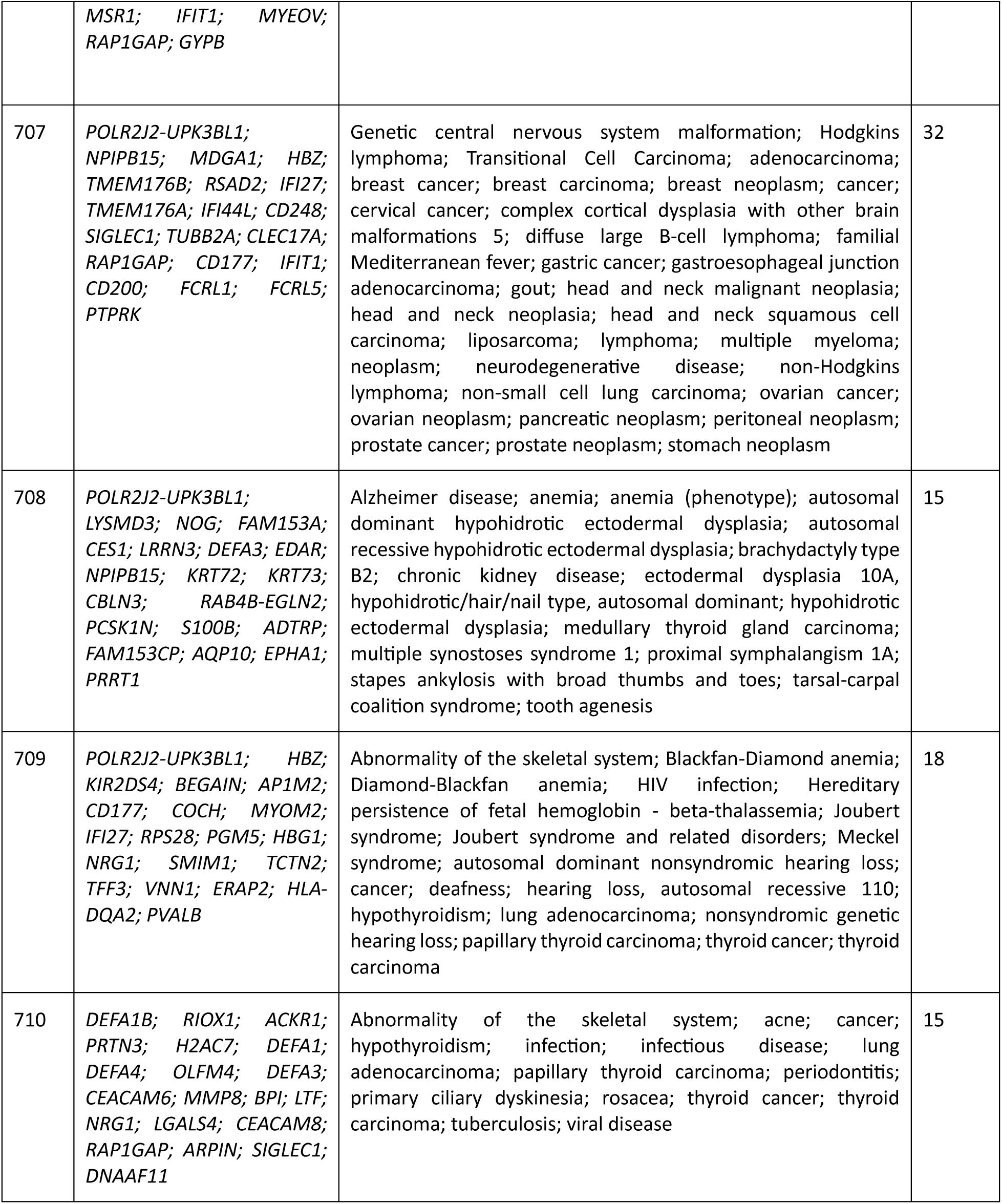
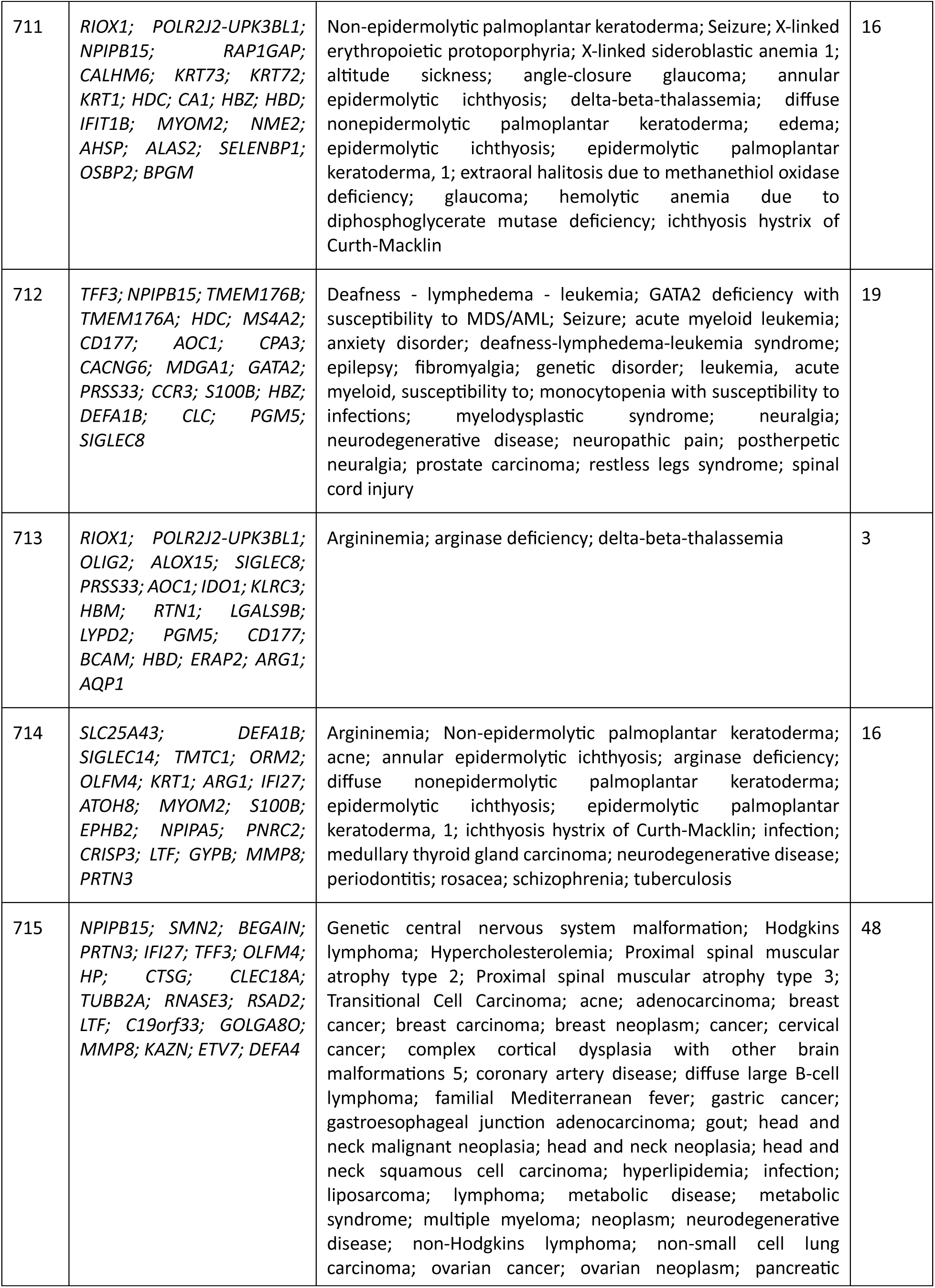
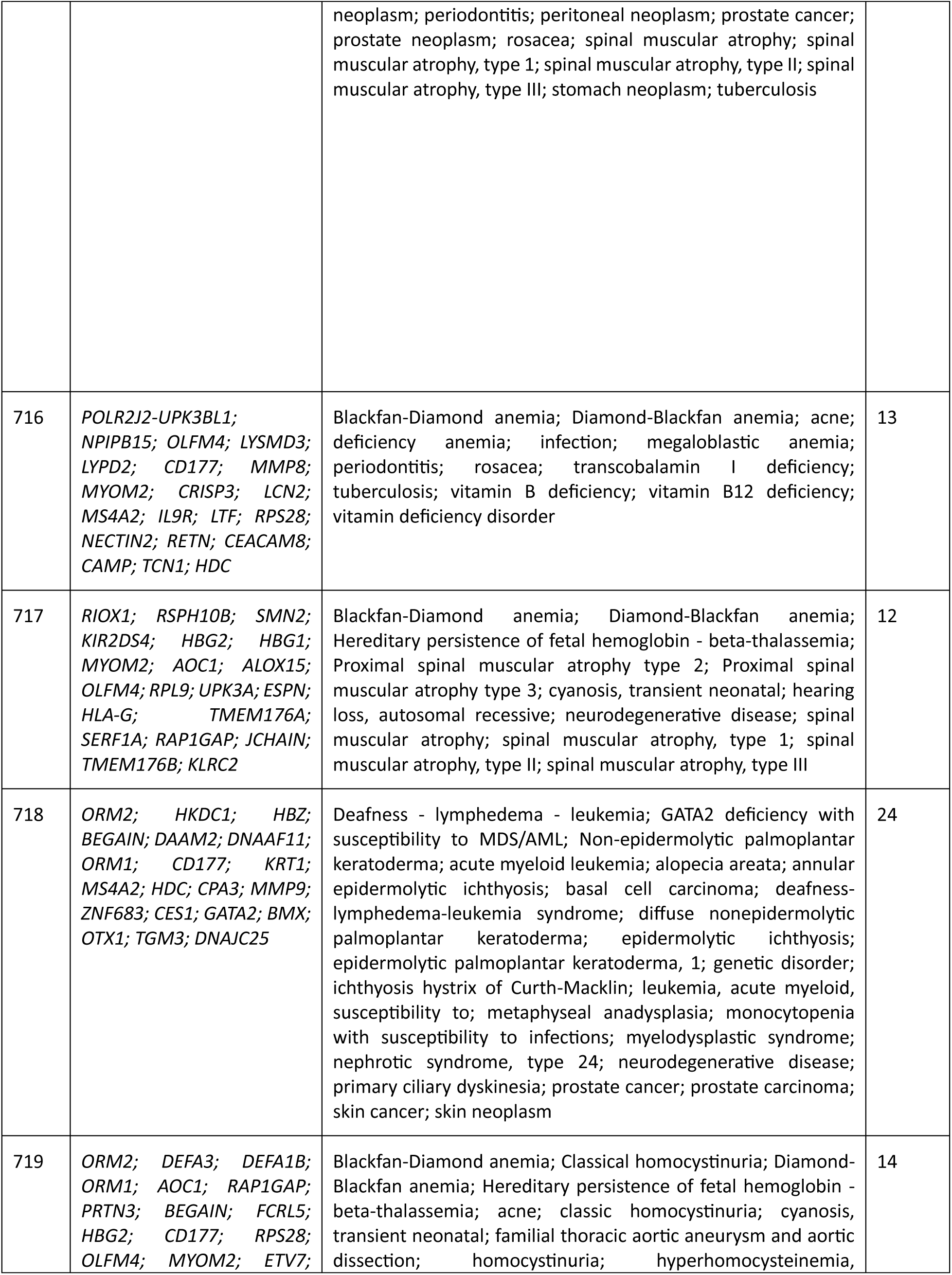
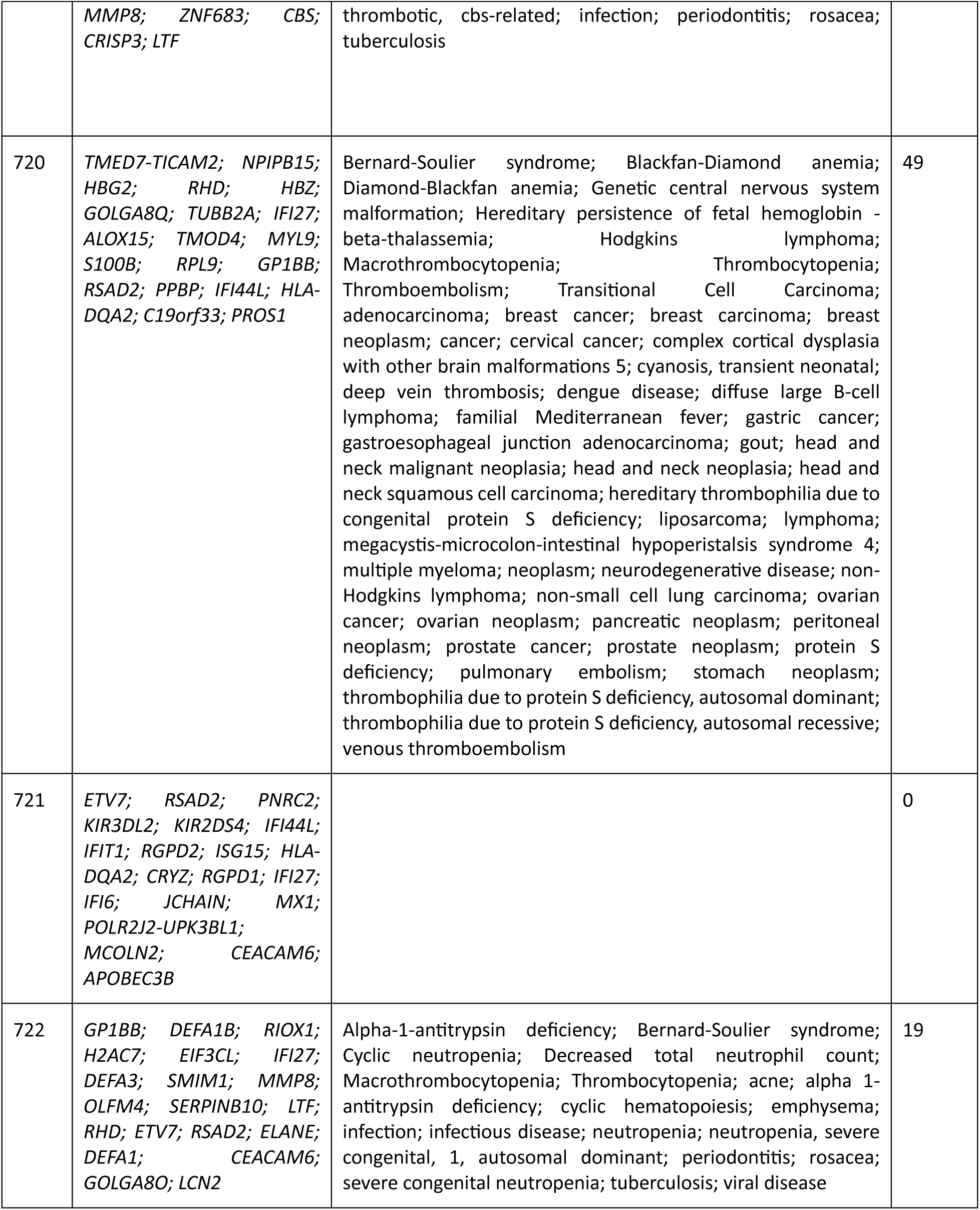
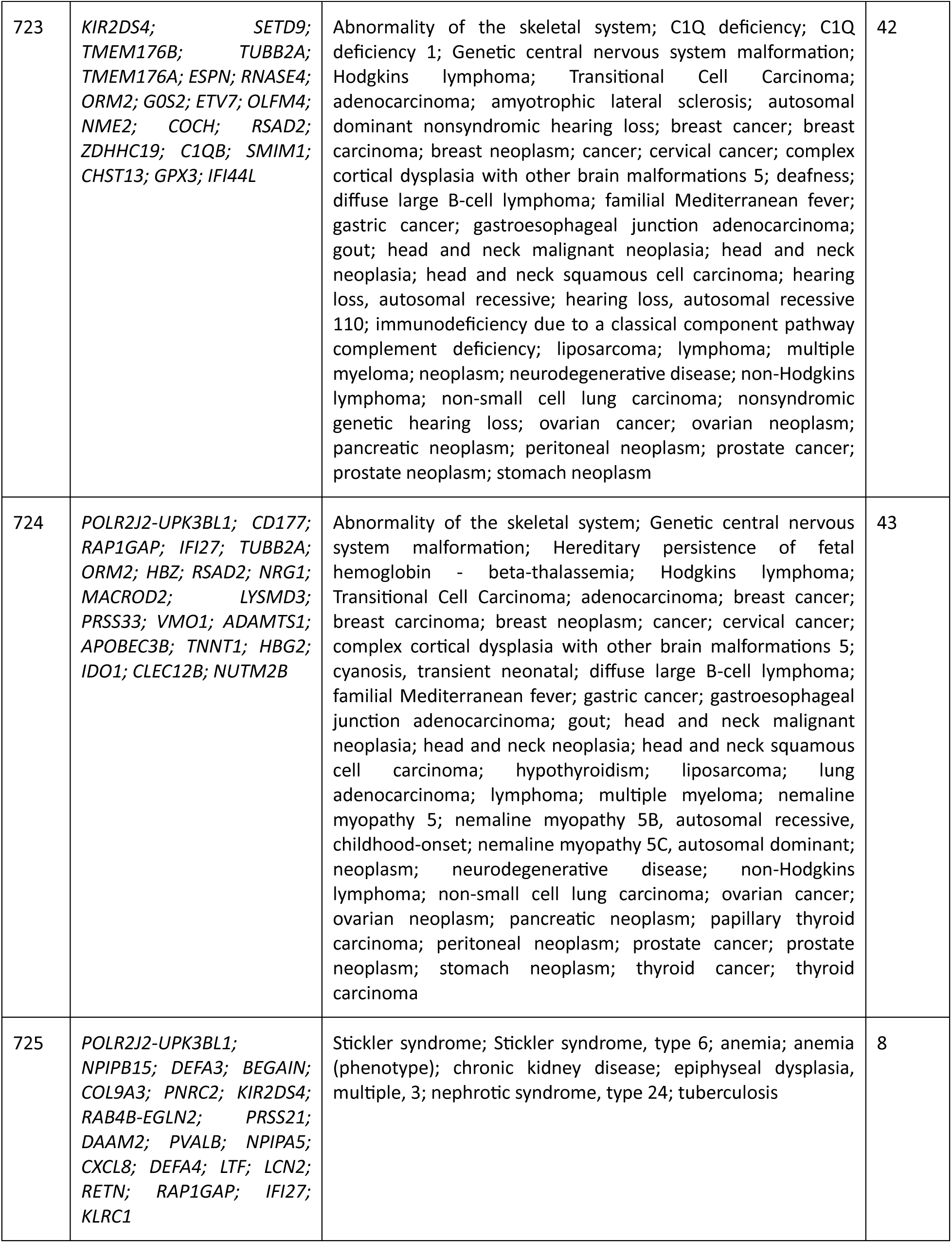
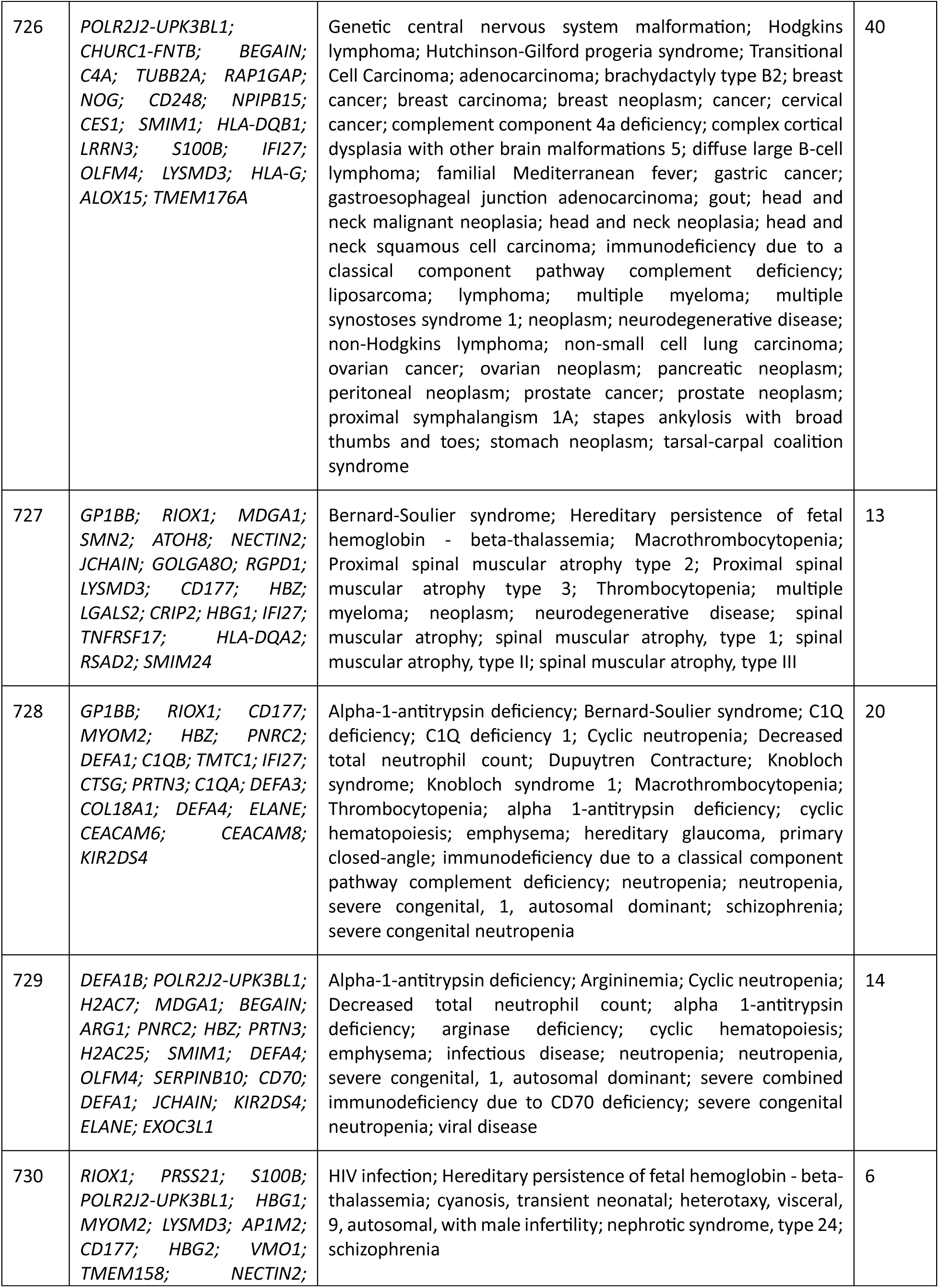
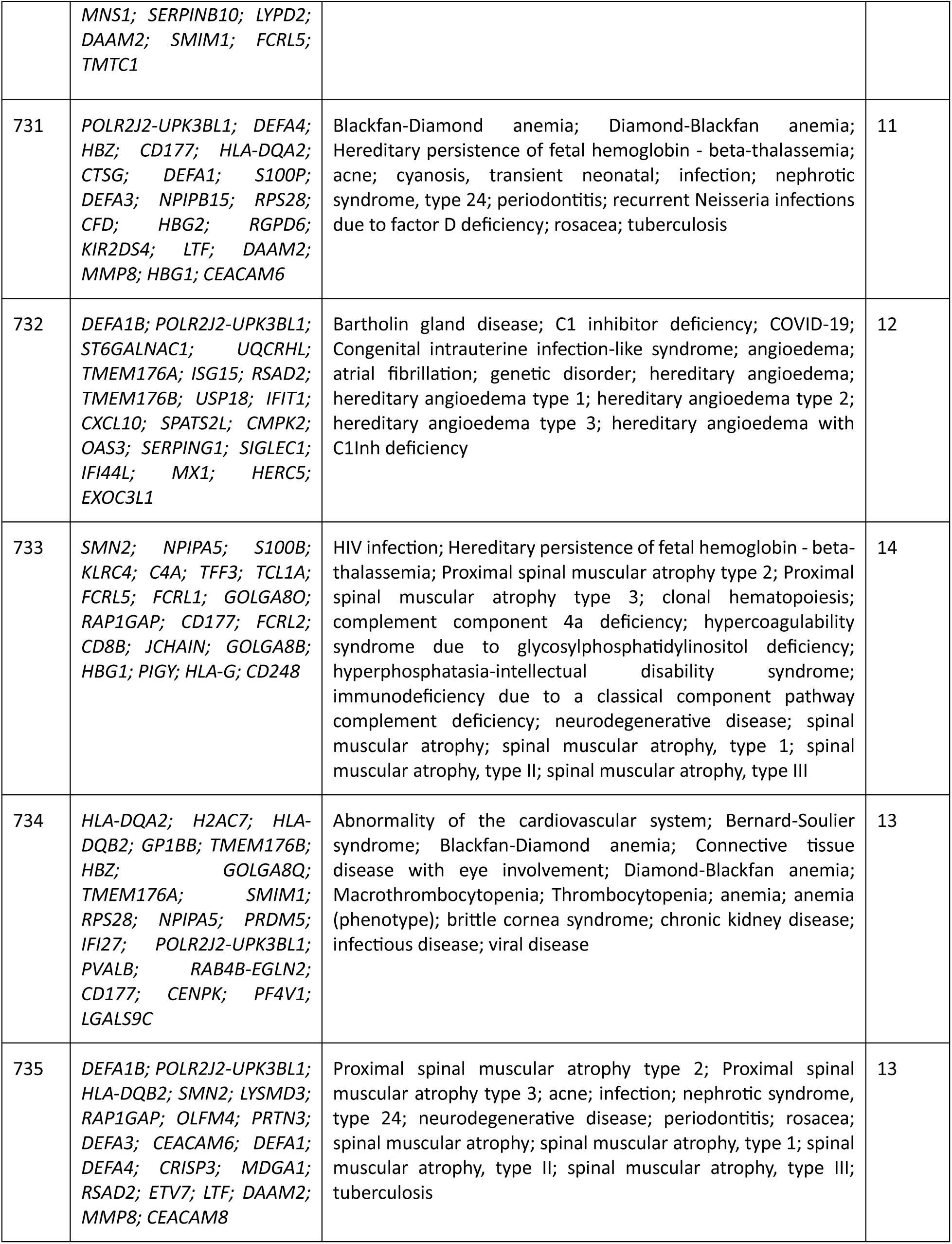
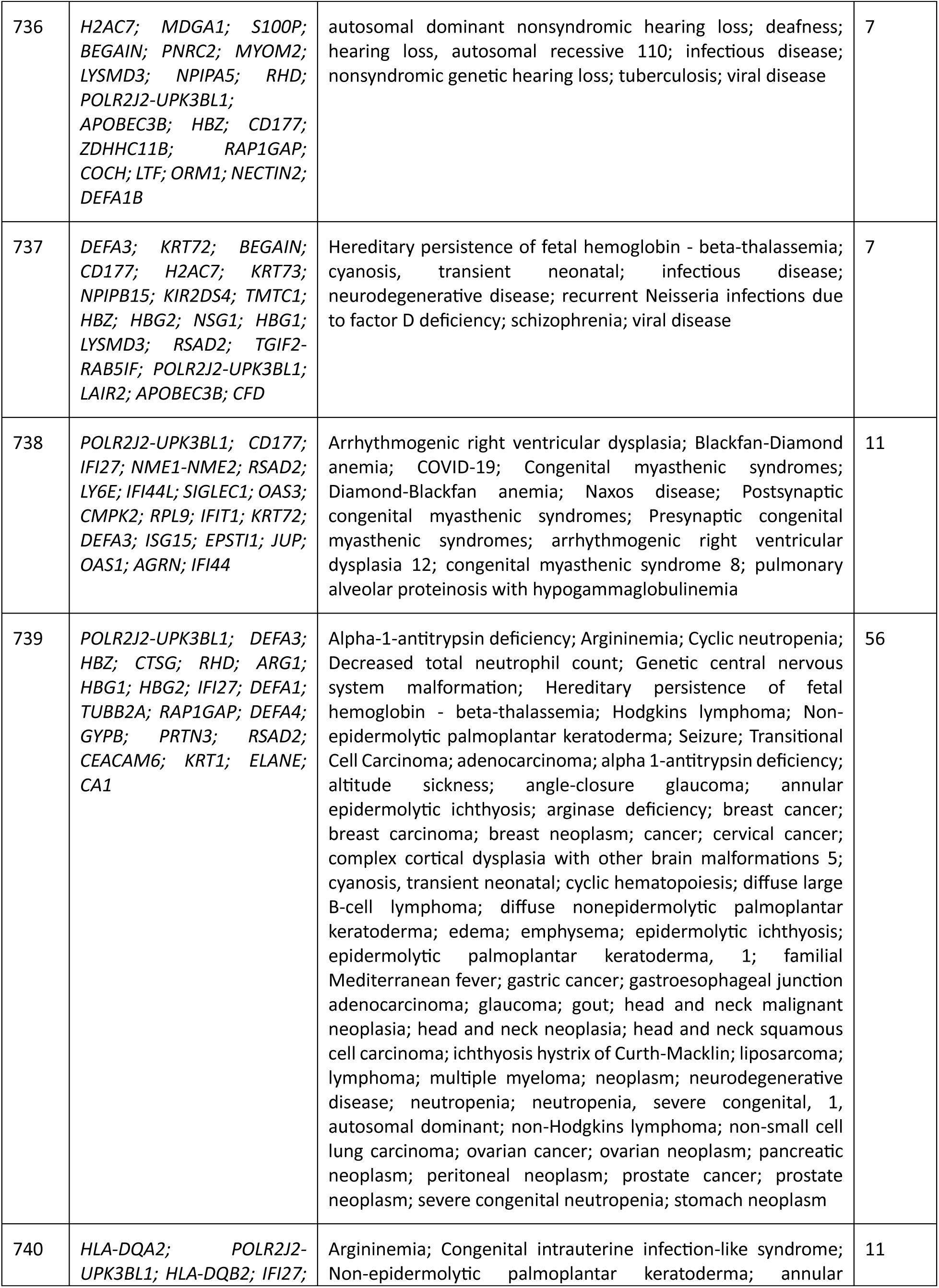
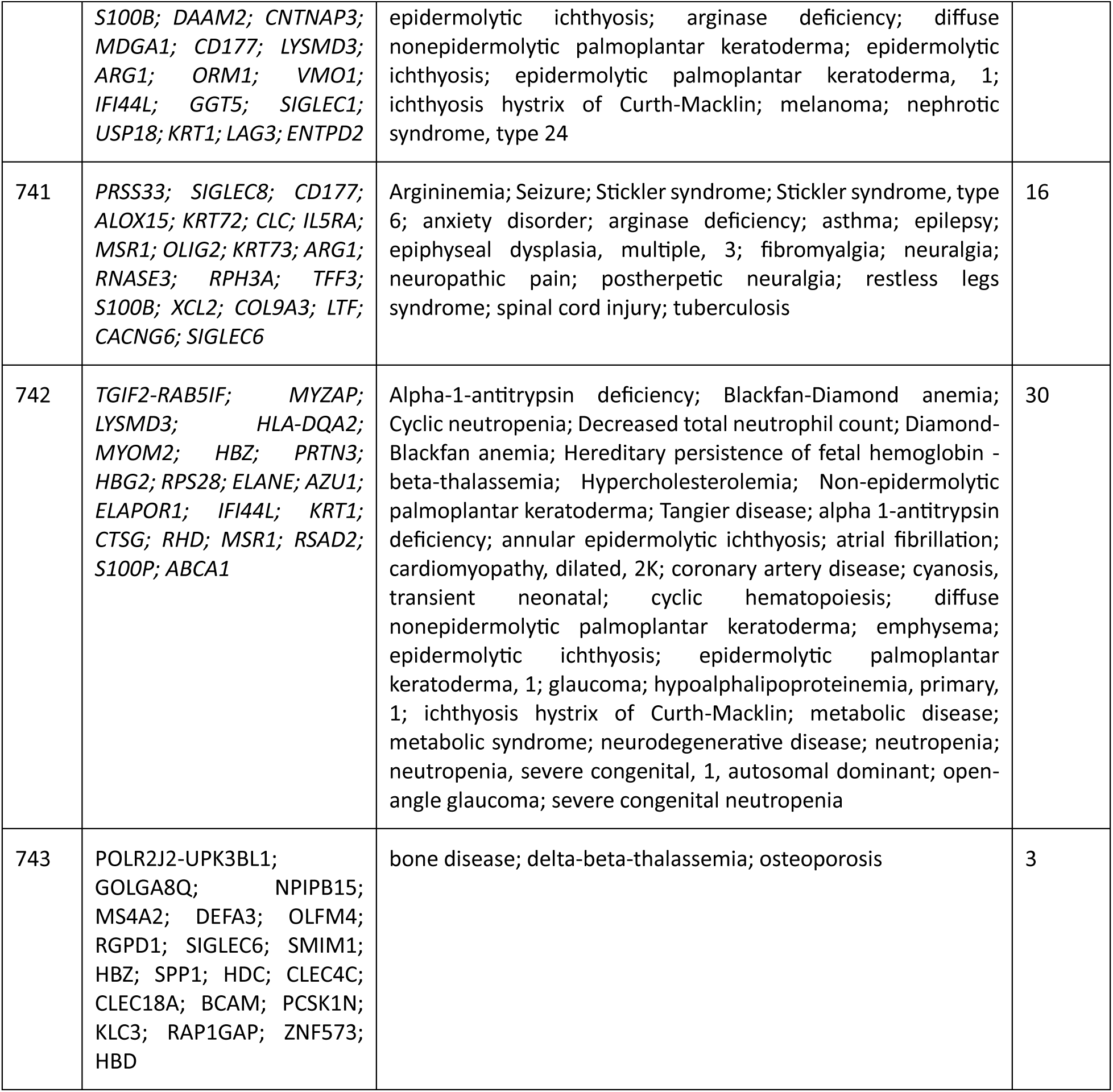
Patient profiles, and gene-disease associations.

## Discussion

This study delves into the application of translational bioinformatics and ML application with multi-omics integration, using a genetically diverse and complex patient cohort. Through constructed individual profiles based on gene expressions, pathogenic variants, and associated diseases, we identified eight mutually predictive genes: *ACKR1, C4B, IFI44L, IFITM3, LGALS2, RETN, RHD,* and *TNFRSF13B.* Next, we applied VAREANT [20] (integrating public annotation databases dbSNP and ClinVar) to identify gene-disease relationships among 148 diseases across RNA-based and WGS cohorts. After manual review and removal of redundancies, 134 distinct diseases were retained. During gene-disease annotation, we realized that there could be a possibility of missing data among annotation databases. Therefore, we conducted an extensive literature review using state-of-the-science studies available from PubMed Central and were able to discover 389 additional diseases associated with these eight genes, which were missing in the annotation databases. Combined, using annotation databases (134 diseases) and scientific literature review (389 diseases), we identified 523 distinct gene-disease associations (Table 2). To better understand the complexities and relationships, we constructed gene-disease networks of all diseases identified through annotation databases (Figure 5A), scientific literature review (Figure 5B), combined annotations from both (Figure 5C), and disease associations with more than one gene only (Figure 5D). Furthermore, we identified that 74 diseases were associated with at least two and at up to seven of the eight mutually predictive genes. These figures reflect original annotation outputs prior to the redundancy removal, therefore include 537 total diseases associated with eight genes.

**Figure 5.**
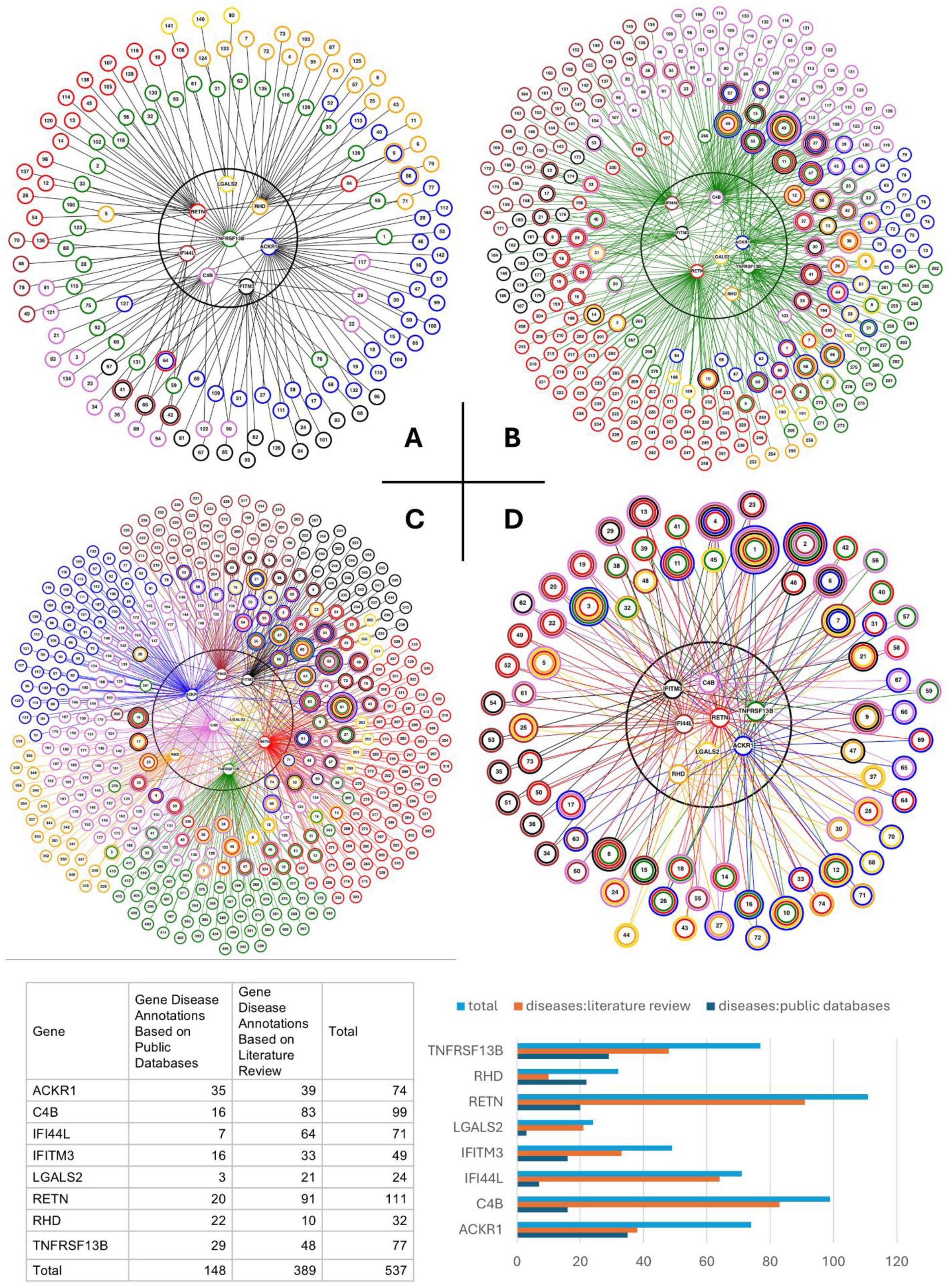
Gene-disease association network. **(A)** Gene-disease association network from the public annotation databases integrated in the methodology. **(B)** Gene-disease association network from the literature review conducted using NIH and PubMed databases. **(C)** Gene-disease associations from both annotation types. **(D)** Gene-disease associations only for diseases that had at least one and at most seven genes overlap of the eight mutually predictive genes.

**Table 2.**
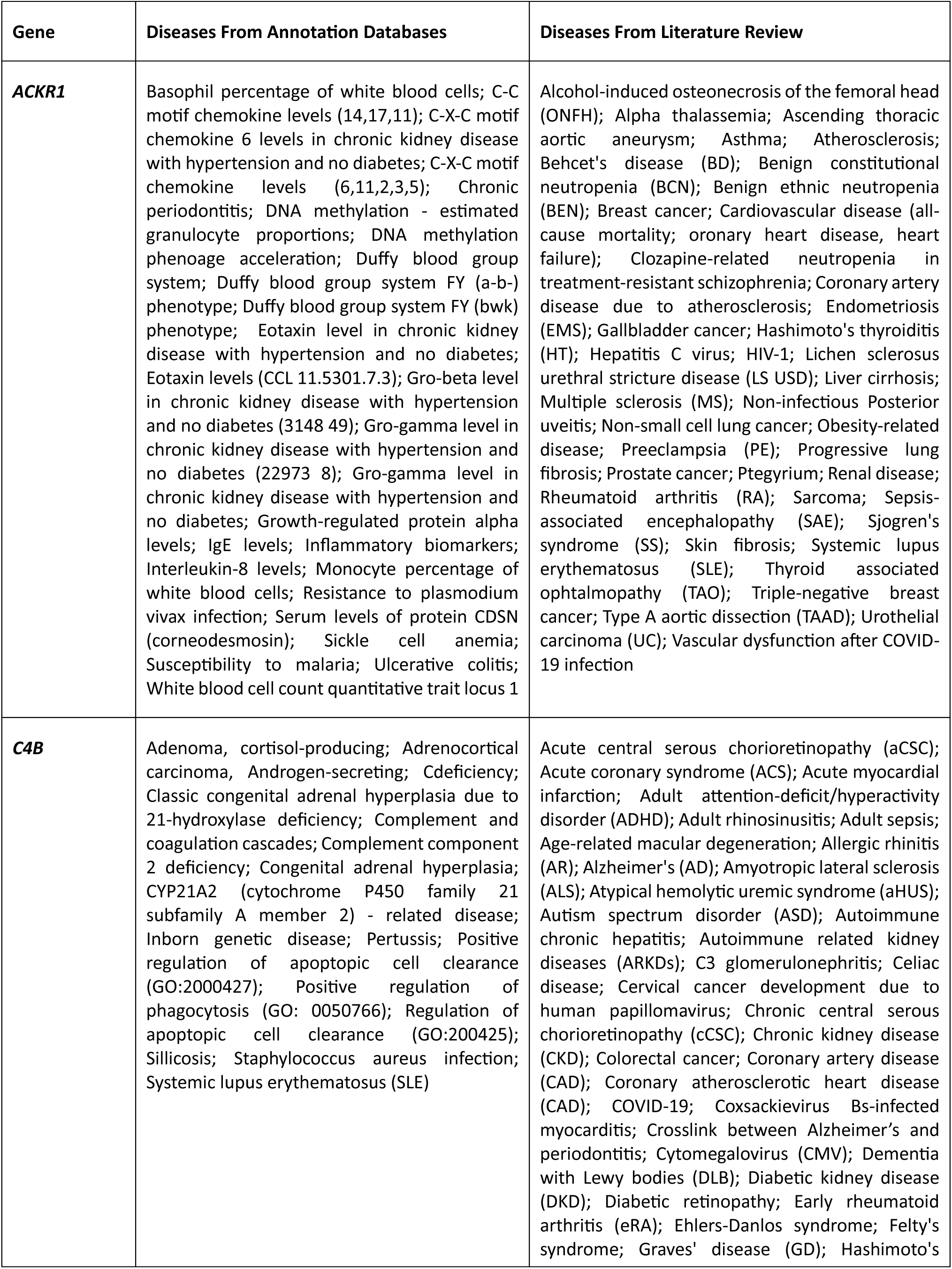
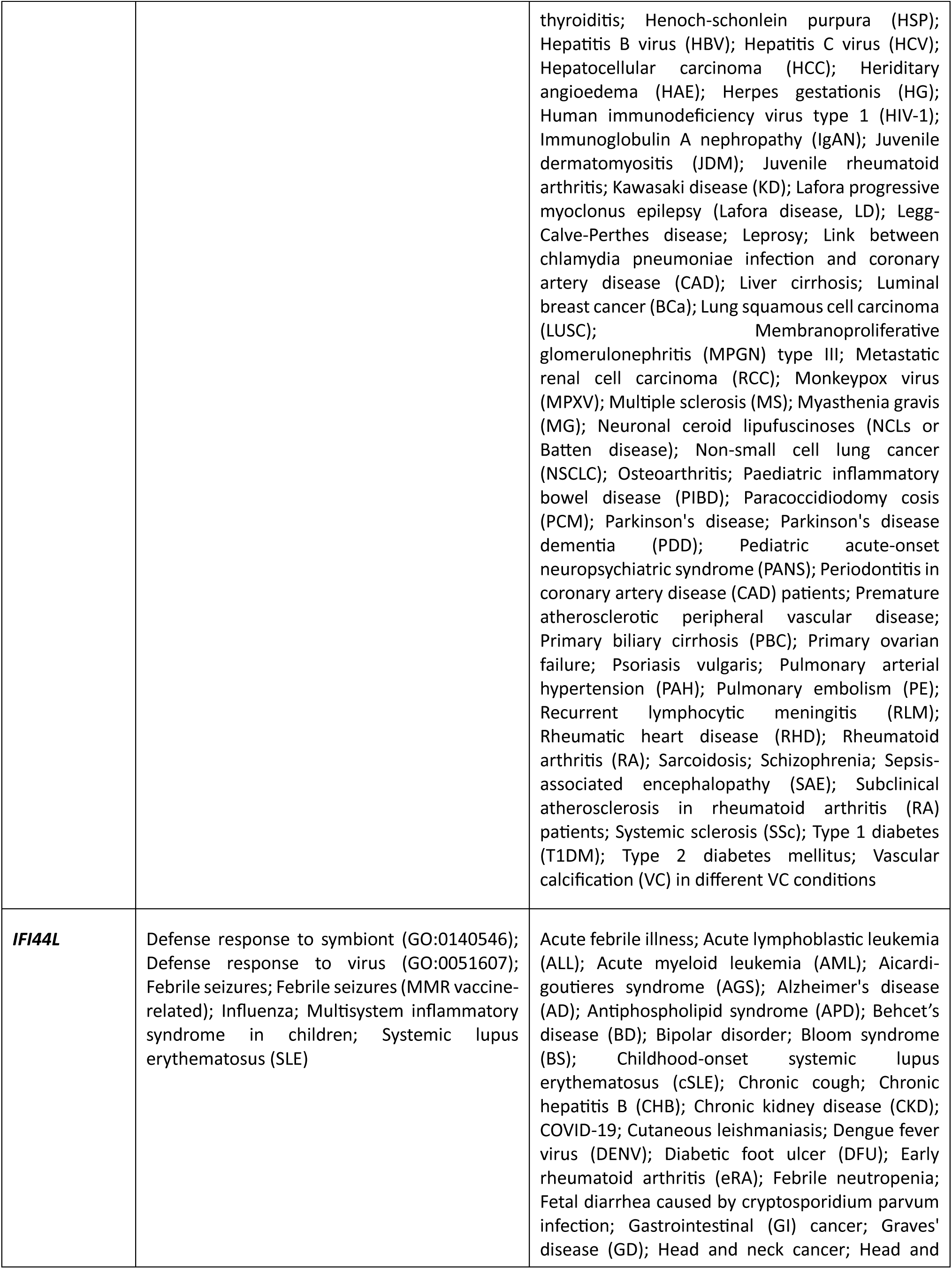
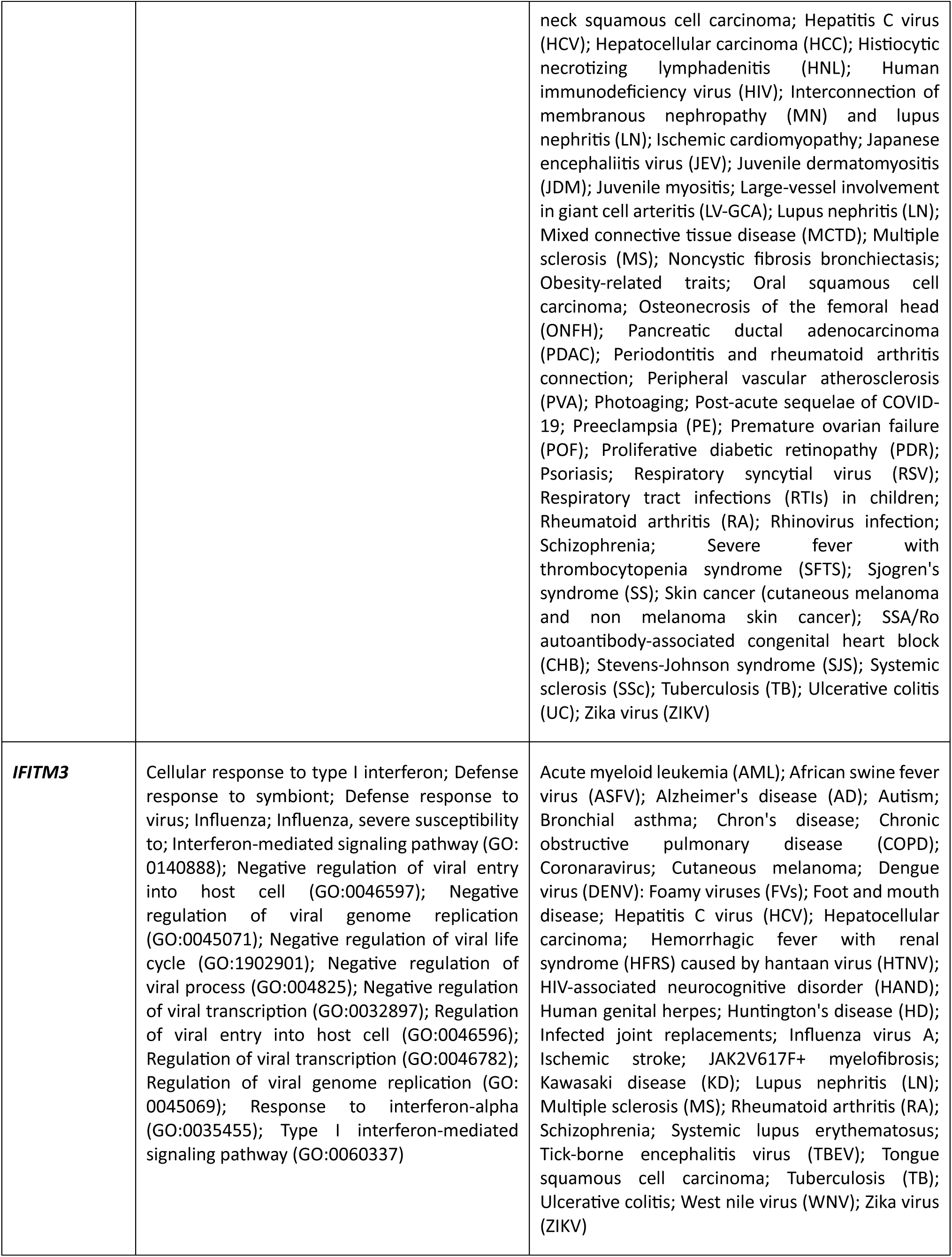
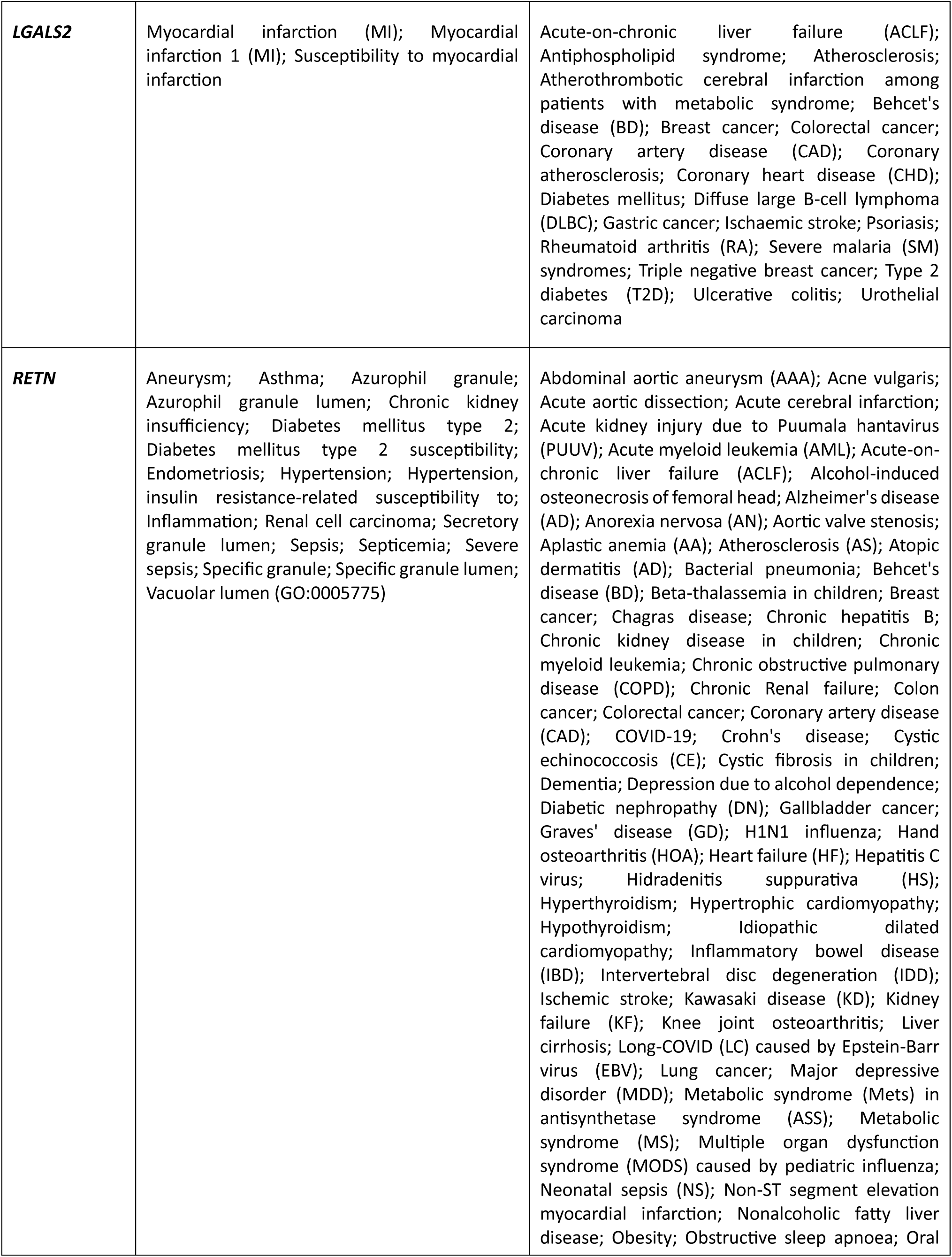
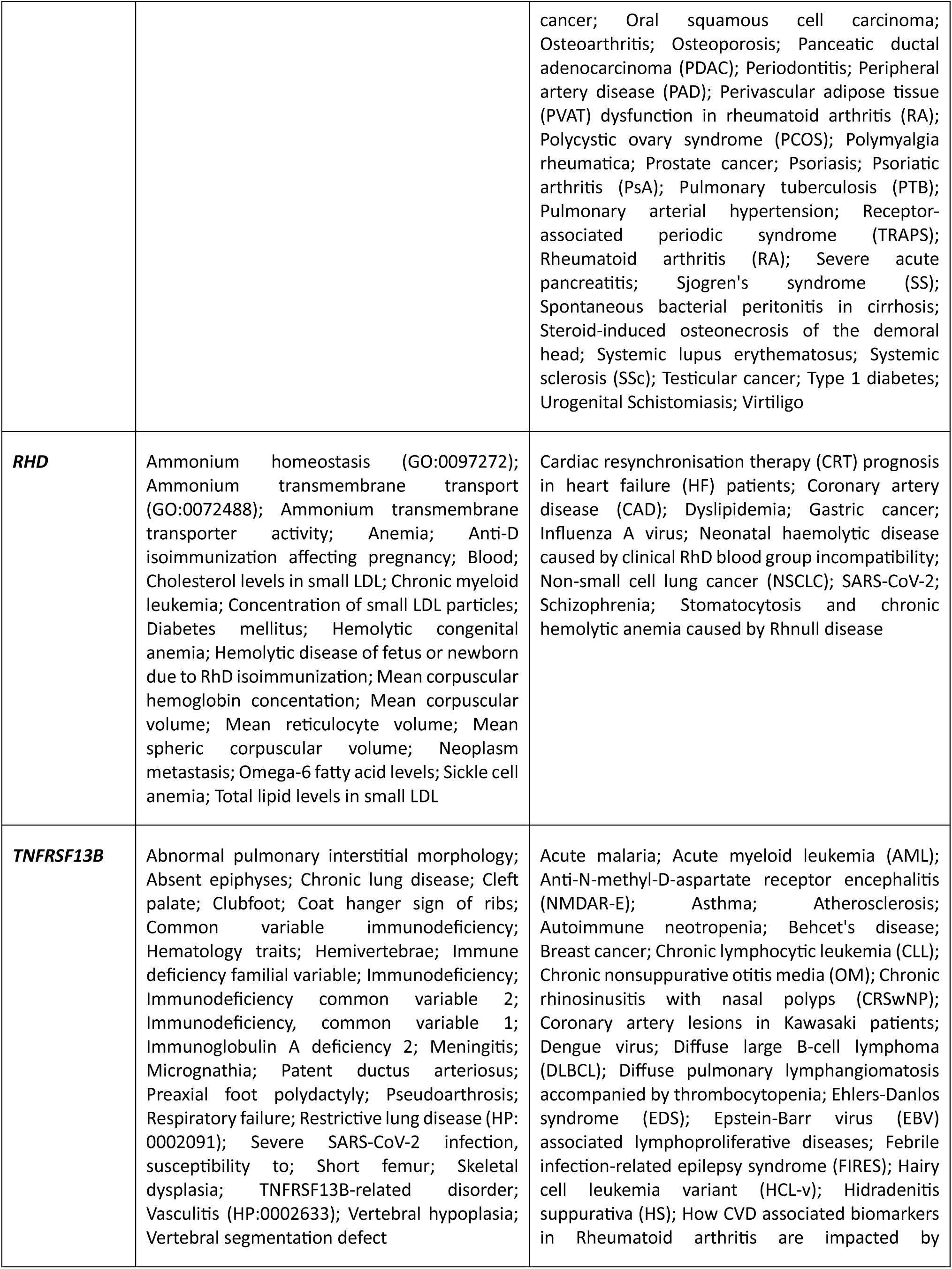
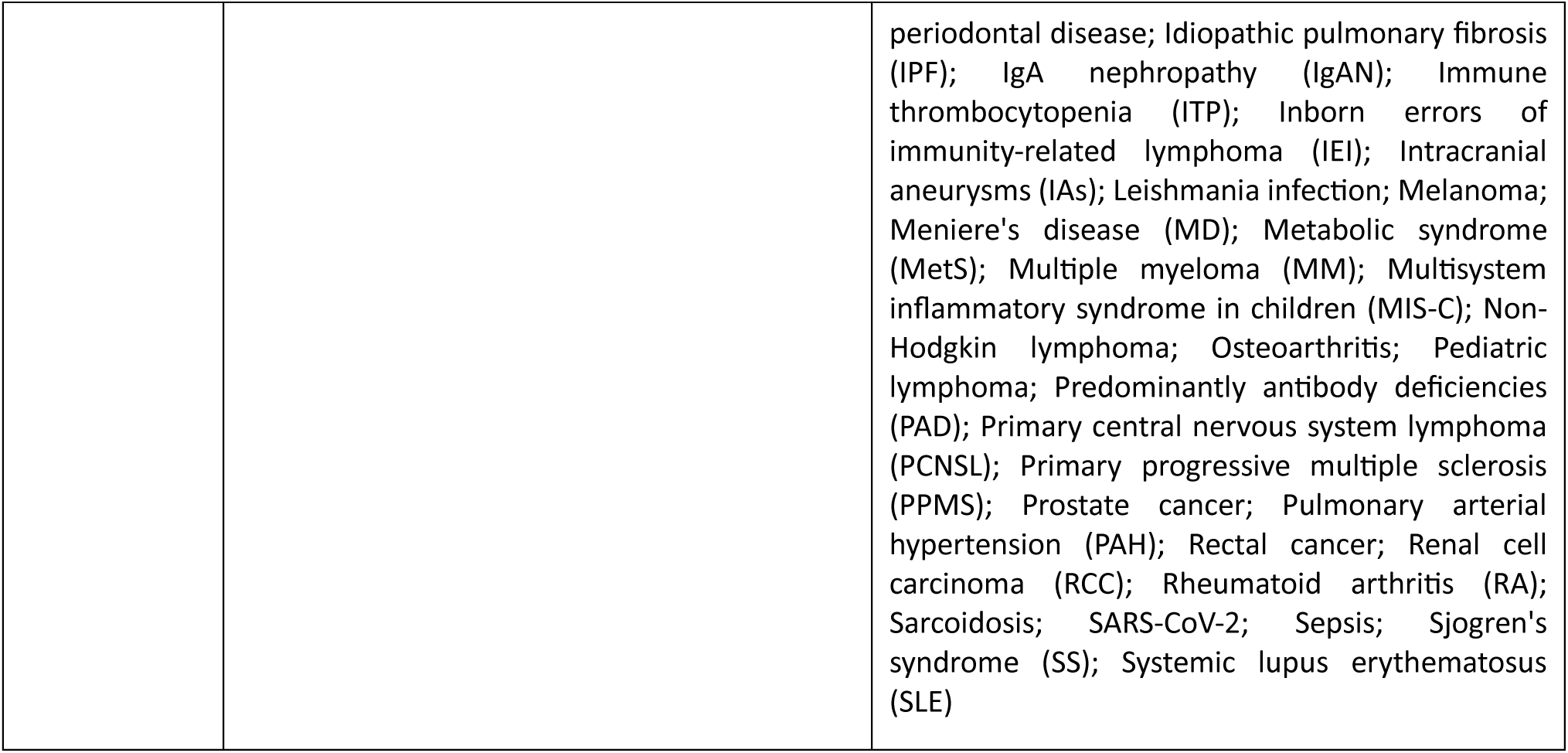
Gene-disease associations derived from annotations databases and discovered through scientific literature review of genes, mutually identified by multi-omics data analyses.

*ACKR1 (*atypical chemokine receptor 1) was found to be one of the eight common genes among patient cohort. It is part of the Duffy blood group and encodes glycosylated membrane receptor protein for malarial parasites Plasmodium vivax and Plasmodium knowlesi [31]. A total of sixty-four diseases were found to be associated with *ACKR1* (twenty-five from annotation databases; and thirty-nine from scientific literature review). Major disease categories associated with this gene include cancer (breast [32] and prostate [33]); CVD and circulatory system disorders (atherosclerosis [34], coronary artery disease [35], and ascending thoracic aortic aneurysm [36]); blood and blood-forming organs diseases (sickle cell anemia [37]); and viral, bacterial, and infectious diseases (malarial disease [38], resistance to plasmodium vivax infection [39], hepatitis C virus [40], and Human Immunodeficiency Virus-1 (HIV-1) [41]). Furthermore, we discovered its binding with other disorders, including musculoskeletal/connective tissue, endocrine/metabolic, neurodevelopmental/nervous system, skin, digestive, respiratory, and rare diseases such as non-infectious posterior uveitis [42] and preeclampsia [43]. Second most common gene among the patient cohort is *C4B* (complement C4B). It encodes complement factor 4, a key effector for adaptive and innate humoral immunity [44]. A total of ninety-nine diseases were found to be associated with C4B (sixteen from annotation databases and eighty-three from scientific literature reviews), which mainly include neurodevelopmental/mental/behavioral and nervous system diseases (schizophrenia [45], attention deficit hyperactivity disorder [46], multiple sclerosis [47], autism [48], Parkinson’s disease [49], and Alzheimer’s [50]); CVD/circulatory systems (myocardial infarction [51]); musculoskeletal system/connective tissue diseases (Systemic Lupus Erythematosus (SLE) [52], Felty’s syndrome [53], juvenile dermatomyositis [54], and rheumatoid arthritis [55]); and endocrine/metabolic disorders (type 1 diabetes mellitus [56], Graves’ disease [57], and congenital adrenal hyperplasia [58]). In addition, we discovered its relationship with diseases e.g., infectious, notably pertussis (whooping cough) [59], cancer (lung [60] and colorectal [61]), six blood and blood-forming organ-related, digestive system and teeth, eye and adnexa, genitourinary system, respiratory system, and miscellaneous diseases such as leprosy [62], Kawasaki disease [63], and amyotrophic lateral sclerosis [64].

*IFI44L* (interferon induced protein 44) is the third most common gene found in the overall patient cohort. It is a protein coding gene that has vast implications in immune response [65] and found to be related to seventy-one diseases (seven from annotation databases, and sixty-four from literature review), which include musculoskeletal/connective tissue disorders (Sjogren’s syndrome [66], juvenile dermatomyositis [67], rheumatoid arthritis [68], and SLE [69]); infectious diseases (HIV [70], COVID-19 [71], and tuberculosis [72]); cancer disorders (hepatocellular carcinoma where *IFI44L* plays a role in cancer stemness, metastasis, and drug resistance [73]); and endocrine/metabolic disorders (Aicardi-Goutieres Syndrome [74]). Additional categories included neurological, skin, blood, CVD/circulatory system, pregnancy/puerperium related, respiratory and rare antiphospholipid syndrome (an autoimmune disorder found in the blood disease category, prominently associated with increased *IFI44L* levels [75]). Fourth most common gene is *IFITM3 (*interferon induced transmembrane protein 3), part of a protein family involved in host defense against pathogens and cellular transformation [76]. A total of forty-nine diseases were found to be associated with *IFITM3* (sixteen from annotation databases and thirty-three from literature review), which mainly include infectious diseases (influenza [77], coronavirus [78], zika virus [79], foot and mouth disease [80], and foamy viruses represented the rarest association [81]); neurological disorders (schizophrenia [82], Alzheimer’s disease [83] and Huntington’s disease [84]); and cancer (hepatocellular carcinoma [85], cutaneous melanoma [86], and *JAK2V617F+* myelofibrosis [87]). However, other associated disease categories include musculoskeletal/connective tissue, respiratory, digestive, and miscellaneous diseases with SLE showing multiple supporting studies [88].

*LGALS2* (galectin 2) found to be the fifth mutual gene in the patient cohort. It is an oxidative stress-response gene largely implicated in cardiovascular and cancer pathways [89]. A total of twenty-four diseases were found to be linked to this gene (three from annotation databases and twenty-one from literature review), which mainly include CVD/circulatory system predominated diseases (myocardial infarction [90] and coronary heart disease [91]); cancer (gastric [92] and colorectal [93]). Furthermore, it showed relationships with musculoskeletal/connective tissue, digestive, endocrine/metabolic disorders, and miscellaneous diseases with Behcet’s disease emerging as one of the only significantly rare diseases [94]. Sixth common gene is *RETN* (resistin), secreted by adipocytes and implicated in metabolic, inflammatory, infectious, and cancer pathways [95]. A total of 110 diseases were identified for *RETN,* (nineteen diseases from annotation databases, and ninety-one from literature review), which mainly include relationships with musculoskeletal/connective tissue diseases (psoriatic arthritis [96], osteoarthritis [97], SLE [98], systemic sclerosis [99], and rheumatoid arthritis [100]); multiple CVD/circulatory system diseases (coronary artery disease [101], atherosclerosis [102], heart failure [103], pulmonary arterial hypertension [105] and the rare Kawasaki disease [101]); cancer (breast [106], colorectal [107], and the rare type of gallbladder [108]); endocrine/metabolic disorders (obesity [109], metabolic syndrome [110], and type 2 diabetes mellitus [111]); infectious diseases (urogenital Schistosomiasis [112]). Additional associations include digestive, neurological, genitourinary, skin, respiratory, blood, and miscellaneous diseases (chronic kidney disease [113], acne vulgaris [114], and Alzheimer’s disease [115]).

*RHD (*Rh blood group D antigen) is seventh most common gene, which is part of the Rh blood group system, which is the most important protein-based blood group system [116]. A total of thirty diseases were found for *RHD* (twenty from annotation databases and ten from literature review). This gene is primarily implicated in anti-D alloimmunization during pregnancy, leading to hemolytic disease of the fetus and newborn anemia [117, 118]. Its additional associations include CVD/circulatory system (coronary artery disease [119]), blood, cancers, infectious and miscellaneous disorders (sickle cell disease [120], chronic myeloid leukemia [121], and schizophrenia [122]), and perinatal period. Interestingly, the Rh-null blood type, also known as “golden blood”, is the rarest blood type in the world, and pertains to diseases such as Rhnull disease, characterized by stomatocytosis and chronic hemolytic anemia [123]. Last common gene is the *TNFRSF13B* (TNF receptor superfamily member 13B). It encodes transmembrane activator and calcium modulator and cyclophilin ligand interactor (TACI) and plays an instrumental role in humoral immunity [124]. A total of seventy six diseases were identified for *TNFRSF13B* (twenty-eight diseases from annotation databases and 48 diseases from literature review), which mainly include cancer (multiple myeloma [125], chronic lymphocytic leukemia [126], and breast cancer [127]); congenital malformations/chromosomal abnormalities; blood diseases (immunodeficiency [128]); musculoskeletal/connective tissue disorders (SLE [129], rheumatoid arthritis [130] and Sjogren’s syndrome [131]); and infectious diseases (SARS-COV-2 [132]). Other categories include diseases associated with respiratory system, CVD/circulatory system, neurological, ear and mastoid process, and miscellaneous diseases (rare Meniere’s disease [133], and IgA nephropathy [134]).

Extensive literature review and association analysis of overlapping diseases among the eight mutually predictive genes revealed rheumatoid arthritis as the most common disease, associated with seven genes (*ACKR1, C4B, IFI44L, IFITM3, LGALS2, RETN,* and *TNFRSF13B).* SLE followed, associated with 6 genes (*ACKR1, RETN, IFITM3, TNFRSF13B, C4B,* and *IFI44L)* with strong literature support across all associations. Behcet’s disease and hepatitis C virus were found to be connected to five genes (*ACKR1, C4B, IFI44L, IFITM3, RETN*). These overlaps highlight shared disease pathways and suggest potential gene-gene interactions demanding further investigation. To clinically validate and better understand the implications of our results, we matched them with the diagnoses extracted from the electronic health records (EPIC Health System). In total, we discovered and reported 412 disorders (after redundancy removal) and from which 116 diseases were matched with diagnosis reported in the clinical records, which include 18 CVD/circulatory system, 13 endocrine/nutritional and metabolic, 12 cancer, 11 musculoskeletal/connective tissue, 10 digestive system, 9 genitourinary system, 8 infectious, 6 nervous system, 6 respiratory system, 5 skin and subcutaneous tissue diseases, 5 neurodevelopmental, 5 blood, and 3 eye and adnexa diseases, and 3 uncategorized/multiple category diseases/isolated and 2 abnormal clinical disorders. Out of these 116 matched diseases, 87 were completely and 29 partially matched based on reflected umbrella terminology, minor disease progression differences, or ICD9–ICD10 coding discrepancies. The remaining 296 unmatched diseases spanned similar categories, including 34 infectious diseases, 33 cancer, 21 CVD/circulatory system, 21 musculoskeletal/connective tissue, 20 blood and blood-forming organ, 15 respiratory system, 12 endocrine/metabolic, 11 congenital malformations/chromosomal abnormalities, 10 nervous system, 6 skin and subcutaneous tissue, 5 eye/adnexa/ear, 3 digestive system diseases, 3 neurodevelopmental, 3 conditions originating in the perinatal period, and 99 uncategorized/combination of categories/isolated in category. In addition, clinical records confirmed strong overlapping for rheumatoid arthritis, systemic lupus erythematosus, and hepatitis C virus. However, Behcet’s disease did not appear in clinical records for the observed patients suggesting a potentially novel inflammatory pathway identified through multi-omics/genomics analysis. We hypothesize that these unmatched and missing diseases in clinical diagnoses may represent potential novel or preclinical pathways, therefore, require further investigation and validation in the future.

## Methods

The overall implemented methodology of this study includes human blood sample collection for multi-omics data generation and preprocessing, and individual and integrated analysis of RNA-seq and WGS data using translational bioinformatics and data driven ML techniques.

### Multi-omics data generation & preprocessing

RNA-seq and WGS were performed on DNA and RNA extracted from blood samples of randomly consented patients (*n* = 96). The sequencing libraries were prepared by random fragmentation, followed by 5’ and 3’ adapter ligation. Adapter-ligated fragments were then PCR amplified and gel purified. Illumina NovaSeq6000 S4 (2×150bp) was utilized for sequencing with 30X coverage using Illumina compatible libraries (e.g., Nextera DNA Flex and TruSeq Stranded mRNA). All human samples were used in accordance with relevant guidelines and regulations, and all experimental protocols were approved by the Institutional Review Board (IRB). We obtained informed consent from all patients. All procedures performed in studies involving human participants were in accordance with the ethical standards of the institution and the 1964 Helsinki Declaration and its later amendments or comparable ethical standards.

Preprocessing of the raw RNA-seq data was carried out using the GVViZ pipeline, which mainly applies Hierarchical Indexing for Spliced Alignment of Transcripts (HISAT) with Bowtie2 to align the sequences against reference human genome (hg38). Next, RNA by Expectation Maximization (RSEM) was utilized for the quantification and identification of Differentially Expressed Genes (DEG) by aligning reads to reference de novo transcriptome assemblies [135]. Furthermore, we performed gene-disease data annotation with GVViZ to identify, classify, and connect significantly expressed genes to human phenotypes and diseases. The expected RNA-seq counts were formatted into a matrix, where rows corresponded to Subject Identifiers (SIDs) and columns represented features. We then retained only those features that were based on protein-coding genes with a mean Transcripts Per Million (TPM) above 10 across the cohort. To preprocess raw WGS data and extract variants into a usable dataset [136], we applied our pipeline i.e., JWES (Java-based Whole Genome/Exome) [137]. JWES combines various tools, including Burrows-Wheeler Aligner (BWA) for reference alignment [138], Genome Analysis Toolkit (GATK) for variant calling [139], and SnpEff for variant annotation [140]. The final outcome of JWES is a Variant Call Formatted (VCF) file, which automatically parsed and stored into relational databases. We utilized VCF extracted information for further downstream analysis, including variant-level quality filtering, classification of coding variations, identification of functionally relevant alleles, and annotation. Preprocessed data from JWES served as the entry point for our subsequent WGS-based variant investigation using ML techniques.

### RNA-seq data analysis

The RNA-seq data analysis methodology includes expression data preprocessing, variance filtering, disease prediction, clustering, and enrichment analysis (Figure 1). In one of our previous studies, we generated RNA-seq dataset (*n* = 61) from a diverse cohort of patients [141]. We utilized these samples to train an accurate, generalizable model for disease prediction. Our custom scikit-learn pipeline initiated with a K-Nearest Neighbors (KNN) imputer [142], using the five nearest neighbors with distance-weighted contributions to fill in missing values. After imputation analysis, a custom quantile transformer mapped the features to a uniform distribution (*n quantiles* = 40). This normalization strategy enabled our model’s applicability across RNA-seq datasets and minimized the batch effects [143]. Next, we performed feature selection using the minimum Redundancy Maximum Relevance (mRMR) algorithm with a Mutual Information Quotient (MIQ) criterion to retain the most informative features (n=15) [144, 145]. Our pipeline’s primary learner is a Random Forest (RF) classifier, instantiated with trees (n=100) and balanced class weights [146, 27]. We implemented a nested cross-validation scheme to provide a performance estimate and tune our model’s hyperparameters. Our outer loop employed repeated stratified 5-fold cross-validation, with each fold repeated five times, resulting in unique dataset splits (n=25). For each outer split, training fold was subjected to an inner cross-validation procedure, where a randomized search over a space of hyperparameters was conducted. We sampled 10 parameter configurations in each inner search, and the pipeline was refitted for each configuration.

After each inner search, the best parameter settings were identified based on the highest F1 score of the minority class. The scikit-learn pipeline was then retrained on the entire outer training set. Probability calibration was performed on this retrained classifier using CalibratedClassifierCV (*method* = ‘sigmoid’), employing a “prefit” strategy such that the calibration module reused the same outer training data to learn a post-processing function that aligned the predicted probabilities with the empirical outcomes [142]. Our calibrated model was evaluated on the unseen outer testing fold, producing per-fold estimates of the F1 score of the healthy control class, Matthews Correlation Coefficient (MCC), balanced accuracy, and standard accuracy. During each of the outer folds (n=25), we recorded both predicted labels and calibrated predicted probabilities on the external holdout data, ultimately generating sets of performance metrics (n=25) that were aggregated and reported with 95% Confidence Intervals (CIs) derived from 1000-sample bootstrapping. Following the nested cross-validation, entire dataset was used for a final hyperparameter search over the same parameter space, relying on a standard 5-fold cross-validation strategy to identify the most effective pipeline settings. The resulting pipeline was trained on the full data and subjected to sigmoid calibration on all samples. Our prediction pipeline was used on our randomly consented cohort after outliers identified by Principal Component Analysis (PCA) were removed. The features were subsetted to the genes previously selected by mRMR. Calibrated probability estimates were generated for each of the remaining samples, after which a 0.70 probability threshold was used to define “confident” positive classifications.

We merged disease predictions with gene expression features to construct a pseudo-labeled version of our randomly consented cohort’s dataset. Using DESeq2, we normalized RNA-seq raw count values [147, 148] and filtered for highly variable genes. Next, we calculated the Coefficient of Variation (CV) for each feature and retained the top 5000 genes. The dataset was re-normalized from raw counts using the same methodology as this set of features, as DESeq2 normalization is dependent on the composition of the entire dataset [148]. Uniform Manifold Approximation and Projection (UMAP) was used to perform dimensionality reduction [149]. We configured the output to two dimensions and incorporated pseudo-labeled information as a soft constraint (*target metric* = ‘categorical’, *target weight* = 0.05). Clustering was carried out on these UMAP embeddings using HDBSCAN [150, 151]. Minimum cluster size and sample parameters were both set to “3”. Next, we used DESeq2 to identify DEG in each cluster relative to all other samples. We deemed genes with an adjusted *p-value* less than “0.05” and a minimum absolute Log2 Fold Change (LFC) greater than “1.5” as differentially expressed. Genes satisfying these cutoffs for each cluster were subjected to enrichment analysis using GSEApy’s Enrichr module [152, 153].

### WGS data analysis

The analysis of WGS data valuably provided insights into complex diseases and their biological bases [154, 155]. To identify relevant and pathogenic mutations associated with disease, we divided our WGS-based methodology into three phases (Figure 1): variant extraction and annotation; pathogenicity classification; and clustering. We processed WGS data in parallel with RNA-seq driven expression data to independently identify and validate significant biomarkers and their relationship with diseases (if any exists) to construct a comprehensive single-patient profile derived from multi-omics data. Next, we employed a two-stage preprocessing pipeline including variant filtering and annotation to extract functionally significant and pathogenic variants, and to identify disease-related biomarkers. For the consistent and comparable results between both methodologies, we performed WGS-based variant analysis using the same preliminary gene set generated by our RNA-seq data methodology. Specifically, all significantly expressed protein-coding genes identified during RNA-seq data analysis formed the primary filtering criterion for extracting targeted variants. Targeting these specific genes ensured greater consistency and comparability with our RNA-seq methodology, as mutually identified biomarkers better reveal underlying pathways and strengthen confidence in retrieved results. It also aids in minimizing the effects of confounding data by narrowing the scope of analysis to only deleterious and impactful variants. Preprocessed variants were extracted using our pipeline i.e., *VAriant REduction and ANnoTation (VAREANT)* [20], with the rest pruned from downstream analysis (Figure 1, W1). As a means of determining disease associations and relevant gene information, each variant was annotated with VAREANT pipeline using two public annotation databases: the Single Nucleotide Polymorphism Database (dbSNP) [156] and ClinVar [157] (Figure 1 W2). Unknown variants (i.e., those without RS numbers) were discarded from the subsequent analysis.

We focused solely on pathogenic variants and those with known disease correlations to identify significant biomarkers to create unique biological profiles per individual. Specifically, we identified all those variants marked as pathogenic, likely pathogenic, or a risk factor for disease (Figure 1, W3). Those otherwise marked benign or with unknown pathogenicity were excluded from the dataset to reduce the influence of confounding and irrelevant data. Before further scoring, it was essential to first normalize gene-variant counts. Since longer genes are more likely to have reads containing pathogenic variants, we first normalized the variant counts based on gene length to avoid underrepresenting shorter genes (Figure 1, W4). Protein-coding gene lengths were sourced from Ensembl BioMart and used to inversely weigh the prevalence of variants when scoring their importance [158]. Pathogenic variants and corresponding genes were then scored using their normalized prevalence among individuals in our cohort (Figure 1, W5).

We divided patients into clusters, based on the distribution of pathogenic variants to identify disease trends and better understand similarities between individuals among the cohort. Patients with similar pathogenic variants are likely to share biological similarities and disease phenotypes. An AI/ML-ready dataset was constructed to aggregate patient data with gene-specific variant data. Particularly, a matrix indicating the prevalence of pathogenic variants in specific genes and individuals was used to cluster patients with an agglomerative (i.e., bottom-up) hierarchical clustering approach (Figure 1, W6). Individuals with the most similar variant profiles are recursively merged to form higher-order clusters. To visualize and interpret the distribution of these clusters, we employed UMAP in 2D (Figure 1, W6). UMAP is a nonlinear projection technique optimized for preserving the local structure of high-dimensional data [149]. Cluster-specific feature annotations, including relevant genes and diseases, were aggregated into individual patient profiles (Figure 1, W7). These patient profiles offer greater comparability for single-patient data to drive precision medicine in addition to having clinical importance in early diagnosis of disease. For each patient, we identified a set of significant genes alongside correlated diseases determined from ClinVar annotations during pathogenicity analysis. Additionally, we utilized ICD-10 codes to further categorize, label, and interpret diseases.

### Gene-disease association and patient profiling

We normalized raw RNA-seq count data from our cohort of randomly consented patients using the same DESeq2 normalization approach utilized in the previous subsection. Next, we calculated CV across all samples to ensure that outlier expression levels would not affect downstream analysis [160]. Raw counts were not re-normalized during any step in this analysis. We built a pipeline to compare a subject of interest to a weighted-average representative of the cohort. First, the dataset was partitioned into two sets: a vector corresponding to initial and a matrix containing all other samples in the cohort. Next, a KNN approach was employed to estimate the distance for every sample in the cohort. Distances were converted into Gaussian weights by centering a normal distribution on the mean of the observed distances and scaling by their standard deviation. This weighting scheme enabled us to construct the profile by taking a weighted average of the reference expression counts, ultimately producing a single synthetic sample that reflects a smooth, population-level baseline for the target patient. Gene-level differences were quantified by computing the LFC for each gene and adding a small constant to prevent division by zero. LFC values were sorted by their absolute magnitude, and the top 20 genes were selected for further investigation. GSEApy’s Enrichr was again utilized, producing patient-specific annotations based on those 20 genes [152, 153]. For each annotation, enrichment statistics, gene sets, and significance measures were determined for individual patients of the cohort.

We utilized Open Targets Platform API (Application Programming Interface) for gene-disease associations with a confidence score ≥ 0.5 [160] to filter out weaker links. Each disease returned by Open Targets was assigned to one of the eighteen disease categories (Blood and blood-forming organ diseases; Nervous system diseases; Immune system diseases; Genitourinary system diseases; Endocrine, nutritional and metabolic diseases; Congenital and genetic diseases; Skin and subcutaneous tissue diseases; Abnormal clinical findings; Musculoskeletal diseases; Respiratory system diseases; Viral/bacterial/infectious diseases; Circulatory system diseases/CVD; Mental/behavioral/neurodevelopmental diseases; Eye and adnexa diseases; Digestive system diseases; Ear, nose and throat diseases; Cancers; and Injuries and wounds) using a three-layer classification system. The first layer used the therapeutic area tags provided directly by Open Targets (e.g., ‘cardiovascular disease’, ‘cancer or benign tumor’) and mapped them into standardized clinical categories. The second layer applied a medical priority rule for diseases tagged under multiple therapeutic areas, ensuring the most clinically specific category was selected over broader parent categories (e.g., diseases tagged as both ‘nervous system disease’ and ‘disorder of visual system’ were correctly assigned to Eye and Adnexa rather than the broader Nervous System). The third layer applied a curated set of name-based overrides for cases where Open Targets’ tagging was clinically inaccurate, for instance, Alzheimer’s disease was tagged as a psychiatric disorder in Open Targets but was correctly reassigned to Nervous System diseases. In total, over 100 such overrides were applied and validated. Additionally, 25 entries returned by Open Targets were identified as non-disease traits (e.g. ‘alcohol drinking’, ‘hair color’, ‘response to statin’) and were excluded from the categorization. Furthermore, to prevent non-specific cancer associations from inflating cancer disorders, a known limitation of broad gene-disease databases, the cancers flag was restricted exclusively to patients carrying a gene from the OncoKB Cancer Gene Census (a curated list of 389 genes with proven causal roles in cancer) [161].

## Declarations

### Ethical Approval and Consent to participate

Informed consent was obtained from all subjects. All human samples were used in accordance with relevant guidelines and regulations, and all experimental protocols were approved by the Institutional Review Board (IRB). All procedures performed in studies involving human participants were in accordance with the ethical standards of the institution and the 1964 Helsinki Declaration and its later amendments or comparable ethical standards.

### Funding

No funding was received.

### Availability of data and material

The data that supports the findings of this study are available from the corresponding author on reasonable request.

### Competing interests

The Authors declare no Competing Financial or Non-Financial Interests.

## Acknowledgments

This research was conducted with the great support of Department of Medicine, Robert Wood Johnson Medical School (RWJMS); and Rutgers Institute for Health, Health Care Policy, and Aging Research (IFH), Rutgers Health. We express sincere gratitude to the current and former members of Ahmed Lab at Rutgers RWJMS/IFH, and appreciate all colleagues, collaborators and institutions who provided direct and indirect insight and expertise that greatly assisted the research and development of this project, most importantly UMKC School of Medicine and UConn School of Medicine. We acknowledge Office of Advanced Research Computing (OARC) for having access to the High-Performance Computing environment and associated computing resources at Rutgers that have contributed to data analysis.

## Author contributions

Z.A. proposed, led, supervised, and drafted this study. Z.A. performed overall project conceptualization, administration, methodology, investigation, data duration, and required funds and resource allocation. Z.A. generated RNA-seq and WGS data, and performed multi-omics data processing, quality checking, and downstream analysis, including gene expression and variant discovery. P.G., W.D., and R.N., participated in predictive analysis, gene-disease annotation, and visualization. E.P. assisted with validation of results using available clinical data and state-of-science literature. S.Z. provided collaborative support and guided the study. All authors have contributed to writing and approved manuscript for publication.

## Biographical note

Z.A. is the Assistant Professor at the Department of Medicine, Rutgers Robert Wood Johnson Medical School, and Core Faculty Member at the Rutgers Institute for Health, Health Care Policy and Aging Research, Rutgers Health. NJ. P.G., W.D., R.N., and E.P., are the Research Assistants/Students at the Ahmed lab, Rutgers IFH/RWJMS. SZ is the Director of UMKC Lab for Investigating Oncology Networks and Systems (LIONS), and Research Assistant Professor at the Department of Biomedical and Health Informatics, at UMKC School of Medicine.

## Notes

### Competing Interest Statement

The authors have declared no competing interest.

### Author Declarations

Informed consent was obtained from all subjects. All human samples were used in accordance with relevant guidelines and regulations, and all experimental protocols were approved by the Institutional Review Board (IRB) at UConn Health and Rutgers University. All procedures performed in studies involving human participants were in accordance with the ethical standards of the institution and the 1964 Helsinki Declaration and its later amendments or comparable ethical standards.

